# Rare missense variants in the SH3 domain of *PSTPIP1* are associated with hidradenitis suppurativa

**DOI:** 10.1101/2022.07.12.22277558

**Authors:** David J. Morales-Heil, Li Cao, Cheryl Sweeney, Anna Malara, Frank Brown, Milan Anadkat, Jessica Kaffenberger, Benjamin Kaffenberger, Peter Nagele, Brian Kirby, Elisha D.O. Roberson

## Abstract

Hidradenitis suppurativa (HS) is a chronic, debilitating skin disease estimated to affect ∼1% of the population, for which few treatment options are available. Risk factors associated with HS include smoking, obesity, and some high penetrance genetic variants. Some rare families have autosomal dominant inheritance. Previous studies have shown that rare loss-of-function variants in genes of the gamma-secretase complex, particularly *nicastrin*, segregate with autosomal dominant HS in some kindred. However, these gamma-secretase variants do not explain much of the overall genetic risk for HS. We performed targeted DNA sequencing of 21 candidate genes in a cohort of 117 individuals with HS to test for an increased burden of rare genetic variants. Candidates included the genes of the gamma-secretase complex, Notch signaling genes, and *PSTPIP1*, a known risk factor for PAPA syndrome. We discovered two pathogenic loss-of-function variants in *nicastrin* that to the best of our knowledge have not been described in HS before. We did not identify significant enrichment of rare missense variants in any gamma-secretase gene, further supporting that loss-of-function in gamma-secretase genes is not a common risk factor. We did, however, identify a statistically significant enrichment of rare variants in the SH3 domain of *PSTPIP1*. Clinical overlap between HS and *PSTPIP1* syndromic features has been noted clinically. Our data suggest that there is shared genetic risk as well, and highlights the need for further population-scale HS genetic research.

## 1. Introduction

Hidradenitis suppurativa (**HS**) is a chronic, debilitating skin disease. Individuals with HS develop painful, deep-seated inflammatory nodules in skin folds, such as the axillae, groin, and perianal regions. Severe HS involves the development of clusters of non-healing abscesses with associated cord-like scarring. HS is estimated to affect 0.5 - 4% of people in different populations^1-4^. Despite the severity of HS, it remains understudied compared to other chronic skin conditions. It has been recognized for decades that HS has an appreciable genetic risk since it can affect multiple members of the same family, and some families with severe HS exhibit autosomal dominant inheritance^5^. The first major insight into HS genetics came from a study that showed some families with highly penetrant, severe HS had loss-of-function (**LoF**) variants in presenilin enhancer 2 (*PSENEN*), presenilin 1 (*PSEN1*), and nicastrin (*NCSTN*), genes that are all components of the gamma-secretase complex^6^. Subsequent screening of gamma-secretase complex genes in families with autosomal dominant HS led to the identification of additional loss-of-function variants in *NCSTN* and *PSENEN*^7-13^. *The vast majority of the gamma-secretase variants discovered in these families to date have been loss-of-function variants in NCSTN*.

Identification of gamma-secretase variants associated with HS was a key breakthrough. However, it remains unclear how these variants mechanistically drive disease. Gamma-secretase is an intramembrane protease complex that cleaves single-pass type 1 transmembrane proteins, [reviewed in ^14^]. It is most often made up of four proteins – nicastrin, presenilin 1, presenilin enhancer 2, and anterior pharynx-defective 1 (APH1). One of the most studied functions of gamma-secretase is its ability to cleave Notch receptors and thereby regulate the Notch signaling pathway. Notch signaling plays a critical role in the development of skin and hair follicles^15; 16^. HS lesions are thought to develop from an initial hyperkeratosis event that leads to occlusion of the hair follicle^17; 18^. It has been hypothesized that dysregulated Notch signaling drives disease in individuals with loss-of-function gamma-secretase variants. This is a logical hypothesis, but Notch is only one of many gamma-secretase substrates^19^. There currently is little direct molecular or genetic evidence that altered Notch signaling is the primary mechanism for familial HS. In a previous keratinocyte cell line knockdown study, we were able to show that low *nicastrin* expression was associated with increased type 1 interferon signaling in HEK001 keratinocytes, rather than disrupted NOTCH pathway signaling^20^. Low nicastrin could therefore lead to disease via multiple different mechanisms.

Although previous research has established an association of gamma-secretase variants with HS, there is also indirect evidence that other genetic risk factors remain to be discovered. Not all families with autosomal dominant HS have gamma-secretase variants. One study found *NCSTN* variants in only 3 of 14 HS affected families^10^. Other studies have failed to show enrichment of rare gamma-secretase variants in sporadic HS cases. In one example only three *NCSTN* variants were found in 48 unrelated individuals with HS: one missense variant of unknown function, one splice-affecting intronic variant that reduced *NCSTN* expression, and one intronic variant of uncertain significance^21^. Another study sequenced the *NCSTN* gene in 95 unrelated individuals with HS and found only one nonsense variant and one missense variant of uncertain significance^22^. Taken together, the results of these different studies suggest that rare variants in *NCSTN* explain only a small portion of the genetic risk for developing HS. Variants in *PSENEN* and *PSEN1* have been identified in some families with highly penetrant HS as well, but there is a lack of data to address how prevalent *PSENEN, PSEN1*, and *APH1* variants are in HS.

Most published genetic screens of HS patients have been limited to amplicon Sanger sequencing of the exons from a subset of gamma-secretase genes, most commonly *NCSTN*. We wanted to perform a broader genetic screen to 1) examine HS-associated rare variant burden in a broader pool of related candidate genes and 2) determine if some of the burden might be due to uncharacterized structural variants. We opted to screen a set of candidate genes with a regional, fosmid-based capture that provides tiled coverage of targeted regions. We settled on a panel of 21 target genes, which included the gamma-secretase complex components and members of the Notch signaling pathway. We also included the genes encoding two proline/serine/threonine phosphatase-interacting proteins: *PSTPIP1* and *PSTPIP2*. Rare *PSTPIP1* genetic variants have been associated with PAPA syndrome, an autoinflammatory syndrome characterized by the development of pyogenic arthritis, pyoderma gangrenosum (**PG**), and acne conglobota ^23^. There are reports of two syndromes that combine features of PAPA with hidradenitis, termed PASH and PAPASH^24; 25^. PASH patients have hidradenitis suppurativa instead of pyogenic arthritis, and PAPASH patients have all the features of PAPA syndrome plus hidradenitis suppurativa. This clinical overlap made us question whether rare variants in *PSTPIP1* or *PSTPIP2* could be risk factors for sporadic HS.

Using our custom capture panel, we screened the DNA of 117 unrelated individuals with HS and compared the rare variant burden in our cohort to the expected distribution of rare variants based on simulated control cohorts using the Genome Aggregation Database (**gnomAD**)^26^. We discovered two rare loss-of-function variants in *NCSTN* that have not been previously reported in HS patients, further supporting the association between *NCSTN* variants and individuals with HS. Additionally, we found a significant enrichment of rare missense variants in the SH3 domain of *PSTPIP1* in our HS cohort. These results suggest that *PSTPIP1* variants may contribute to the genetic risk for HS in addition to PAPA and PAPA-like syndromes. We also screened for deletions and amplifications using comparative sequencing coverage analysis. We did not identify large deletion or amplification events in gamma-secretase or NOTCH pathway genes as a significant contributor to HS-associated genetic risk factors.

## 2. Results

### 2.1. Study cohort description

We performed targeted capture sequencing on 117 HS patients collected from three different cohorts: 1) a cohort collected through the Dermatology Clinic at St. Vincent’s University Hospital in Dublin, Ireland (**SVUH**; n=70); 2) a cohort collected through the Division of Dermatology at Washington University in St. Louis, Missouri, USA (**WU**; n=25); and 3) a cohort collected through the Division of Dermatology at Ohio State University in Columbus, Ohio, USA (**OSU**; n=22). Informed consent was obtained from all participants prior to enrollment in accord with the Declaration of Helsinki. The patients recruited for our study were predominantly between the ages of 21-50 years old (SVUH 85%; OSU 81%; WU N/A). Most participants identified as women (SVUH 79%; OSU 82%; WU N/A). Of patients who responded to questions regarding family history of disease, approximately 50% self-reported at least one affected relative. Summary information about the cohorts is presented in **Table 1**.

**Table 1.**
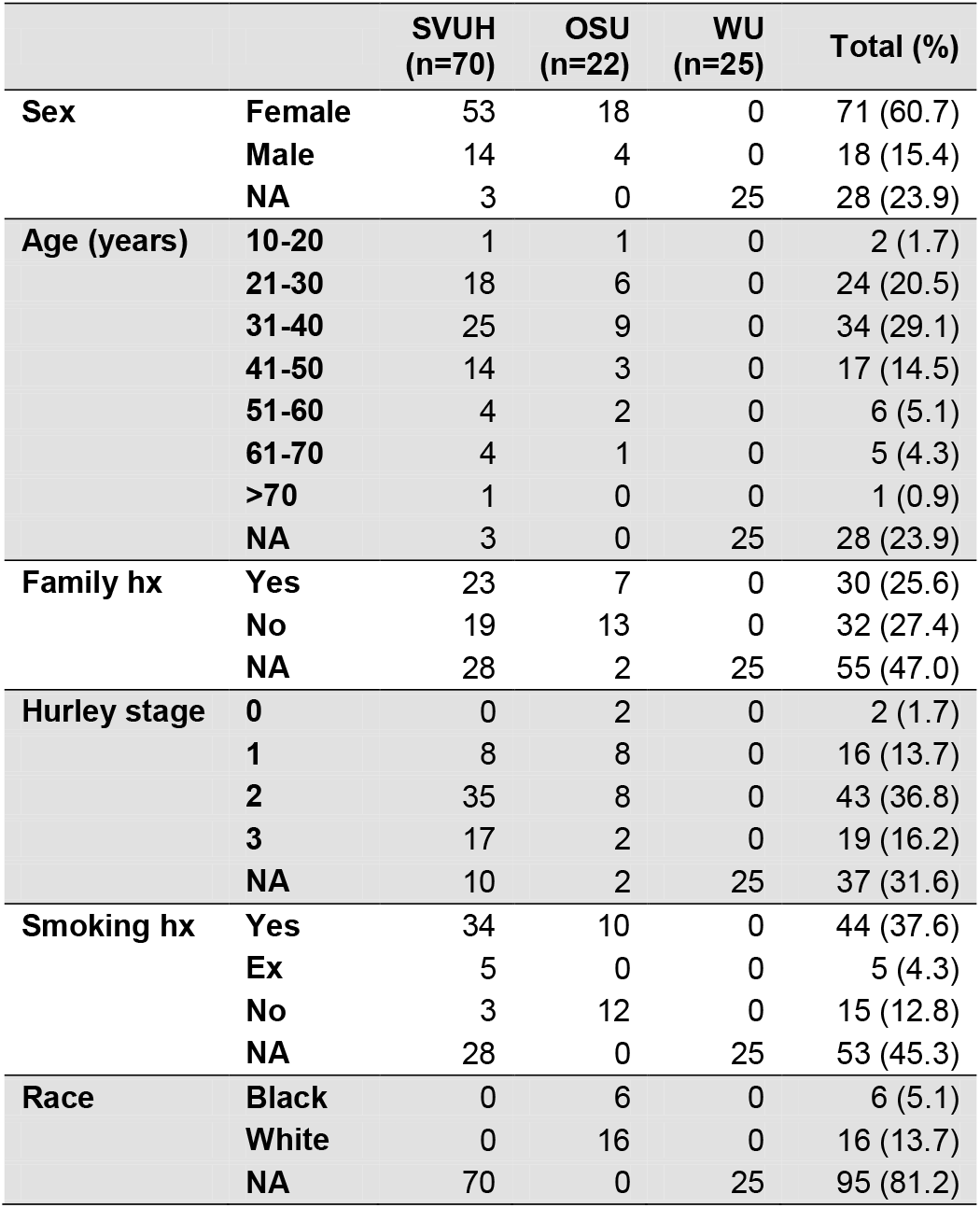
Cohort demographics. Hs patient cohort was collected St. Vincent’s University Hospital (**SVUH**), Ohio State University (**OSU**), and Washington University in St. Louis (**WU**). Not all individuals responded to all survey questions during sample collection, and no demographic information was available for WU samples. The available information reflects known HS epidemiology by affecting more women than men and being associated with smoking. Abbreviations: NA, Not Available; hx, history.

### 2.2. Targeted capture performance

We isolated genomic DNA from patient saliva or blood and performed targeted capture sequencing using capture probes generated from fosmids and bacterial artificial chromosomes (**BACs**) containing human genomic sequence, as previously described^27^. Our capture panel used 65 fosmids or BACs to capture 21 non-overlapping regions in the genome that covered a total of ∼2.5Mb (**Table ST1**). The capture targets included 21 genes of interest that can be summarized as gamma-secretase genes (*PSEN1, PSEN2, APH1A, APH1B, NCSTN, PSENEN*), Notch receptors (*NOTCH1, NOTCH2, NOTCH3, NOTCH4*), Notch transcriptional regulators (*MAML1, MAML2, MAML3, RBPJ*), Notch ligands (*JAG1, JAG2, DLL1, DLL3, DLL4*), and other genes of interest (*PSTPIP1, PSTPIP2*). The captured patient DNAs were sequenced to approximately ∼100X mean coverage per individual (**Figure S1**).

We called variant impacts with respect to one chosen isoform of each gene (**Table ST2** and description in Methods). We identified a total of 16,764 genetic variants in the captured regions of our patient cohort. Of these, 10,901 occurred in gene-associated regions, defined as the regions between the first and last exonic bases of our genes of interest (**Table ST8**). The vast majority of gene-associated variants (10,622) were non-coding (UTR and intronic) variants. We identified 159 synonymous and 120 missense/nonsense rare variants within the coding regions of our targeted genes (**Tables ST3-ST5**).

### 2.3. Approach to rare variant burden tests without a control cohort

Aside from a handful of small polyQ tract indels in *MAML2* and *MAML3*, the other protein-affecting variants that we observed consisted of missense single nucleotide variants of uncertain significance and one single nucleotide deletion. We, therefore, wanted to test for an increased burden of rare protein-affecting variants in our HS patients. While we could have collected a similarly sized control cohort, we instead relied on rare variant information from the Genome Aggregation Database (**gnomAD**). This decision was mainly motivated by the fact that this database has rare variant frequency information from over a hundred thousand individuals. While not all controls, they are not enriched for HS patients and give us a better view of the range of tolerable protein-affecting variation in our candidate genes. A proof-of-principle of this kind of approach has been previously published for a cohort of individuals with idiopathic hypogonadotropic hypogonadism, which identified variant enrichments in genes previously known to be relevant to the disease^28^.

Whether or not an allele is over-represented in a cohort depends upon the expected alternate allele frequency for the population under examination. We didn’t have self-reported race or ethnicity for all individuals in this cohort, as it was assembled from several different groups with varying levels of detail. Those that did report race categorized themselves as Black or White. However, race as a categorical classification is a social construct. Systematic differences in genetic variant allele frequencies represent historical geographic isolation of populations which may or may not accurately reflect how an individual identifies socially. gnomAD contains human genetic information derived from ∼140,000 individuals of various geographic ancestries. We leveraged this data from gnomAD to identify which of their populations our samples most resembled for the overall capture region.

We extracted all variants from the gnomAD genome and exome datasets that fell within our targeted capture region. These variants, and our targeted capture variants, were then filtered through a quality control (**QC**) process, compiled, and annotated (see Methods). We used the non-Finnish European (**NFE**), African/African American (**AFR**), Admixed American/Latino (**AMR**), East Asian (**EAS**), and South Asian (**SAS**) subpopulation alternate allele frequencies from gnomAD to simulate genotypes for 1000 individuals for each subpopulation over our targeted capture regions. Principal Components Analysis (**PCA**) of these simulated genotypes clustered by geographic ancestry (**Fig. 1a-c**). Given that our capture region was a small fraction of the genome, some populations were more difficult to distinguish than others. For example, NFE and SAS somewhat overlapped with each other.

**Figure 1.**
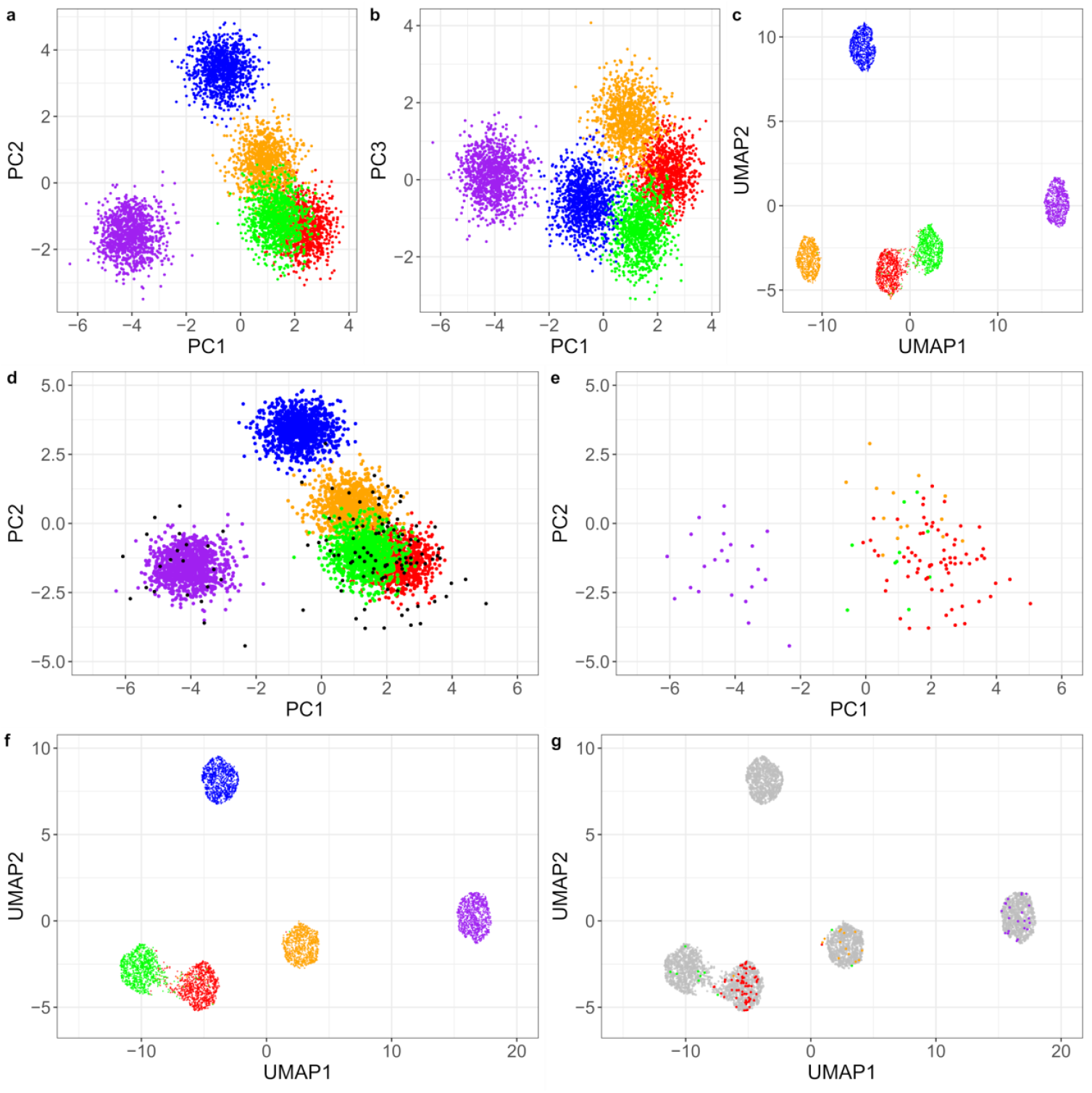
Ancestry categorization of the patient cohort. Data for simulated controls (1000 individuals / population) and our cohort (n=117). **a**. The PC1 / PC2 control plot shows the separation between African and non-African ancestries. **b**. The other ancestries separate more from each other on PC1 and PC3, though there is still substantial overlap. **c**. PCA-informed UMAP projections of controls have better spatial separation between ancestries than the PCA alone **d**. The PC1 / PC2 projection with our cohort overlaid (black dots). Our samples correspond well to the gnomAD populations. **e**. The same projection with our samples alone, colored by assigned ancestry. **f**. PCA-informed UMAP of controls when our samples are included. **g**. The same projection with our cohort alone, colored by ancestry assignment. Ancestries: African / African American, purple; Admixed American / Latino, yellow; East Asian, blue; Non-Finnish European, red; South Asian, green. Abbreviations: PC, principal component; UMAP, uniform manifold approximation and projection.

We next repeated the PCA analysis and included our HS cohort genotypes. Using the simulated genotypes as a training set, we performed k-nearest neighbor (**k-NN**) analysis to categorize our patients into the gnomAD subpopulation ancestries that they most closely resembled (**Fig. 1d-g**). This categorized our cohort as being made up of 71 non-Finnish European individuals, 22 African American individuals, 14 Latino/Admixed American individuals, and 10 South Asian individuals. It’s worth noting that this method forced a categorization into a single population group, and simply reflects the closest matching gnomAD population. We only had self-reported race information for one of our patient cohorts. Our method classified all six individuals that self-identified as Black into the simulated African/African-American cluster. The 16 other individuals for whom we had self-declared race identified as White. Our method classified 13 of these individuals as non-Finnish European, 2 as Latino/Admixed American, and one as South Asian. These disagreements with self-declared ancestry could reflect the limited classification power of our capture region, ancestry unknown to the individual, or a combination of these factors.

Identifying the nearest matching gnomAD ancestry subpopulation for each patient in our cohort provided us with a way to subsequently simulate the number of expected rare (minor allele frequency <1%) missense variants in an ancestry-matched control cohort (**Fig. 2**). Briefly, we classified each gnomAD QC pass variant as either circulating in the population or a singleton, which we defined as a variant observed no more than one time in all population groups combined. Approximately 50% of the gnomAD rare protein-affecting variants that were extracted for our analysis were singletons (**Fig. S2**). This observation is consistent with the proportion of variants that were observed to be singletons in the Exome Aggregation Consortium (**ExAC**)^29^. To simulate genotypes for a control cohort, we handled circulating variants and singletons differently. For each circulating variant, we drew two random numbers to simulate two alleles at the position, and the individual was credited with a rare variant for each random number that was less than the minor allele frequency for the variant. For singletons, we calculated gene-specific singleton rates. These gene-specific singleton frequencies were similar between most subpopulation groups, and there was a clear linear relationship between singleton rate and gene length (**Fig. S2**). After simulating all circulating variants we again drew two random numbers. The individual was credited with a singleton allele in that gene for each number that was less than the gene-specific singleton rate. For each simulated cohort, we counted how many rare protein-affecting variants were observed per gene, and repeated this process for 200 cohort simulations to estimate the average number of rare protein-affecting variants to expect for each gene. We then had to test whether the number of rare protein affecting variants in our HS cohort was greater than the expectation in controls. We tested for enrichment of rare variants in each gene using a Poisson distribution with the average number of simulated rare variants as the expected lambda parameter, correcting the p-values by the Bonferroni method. Genes with several expected variants most clearly followed a Poisson distribution (**Fig. S3**).

**Figure 2.**
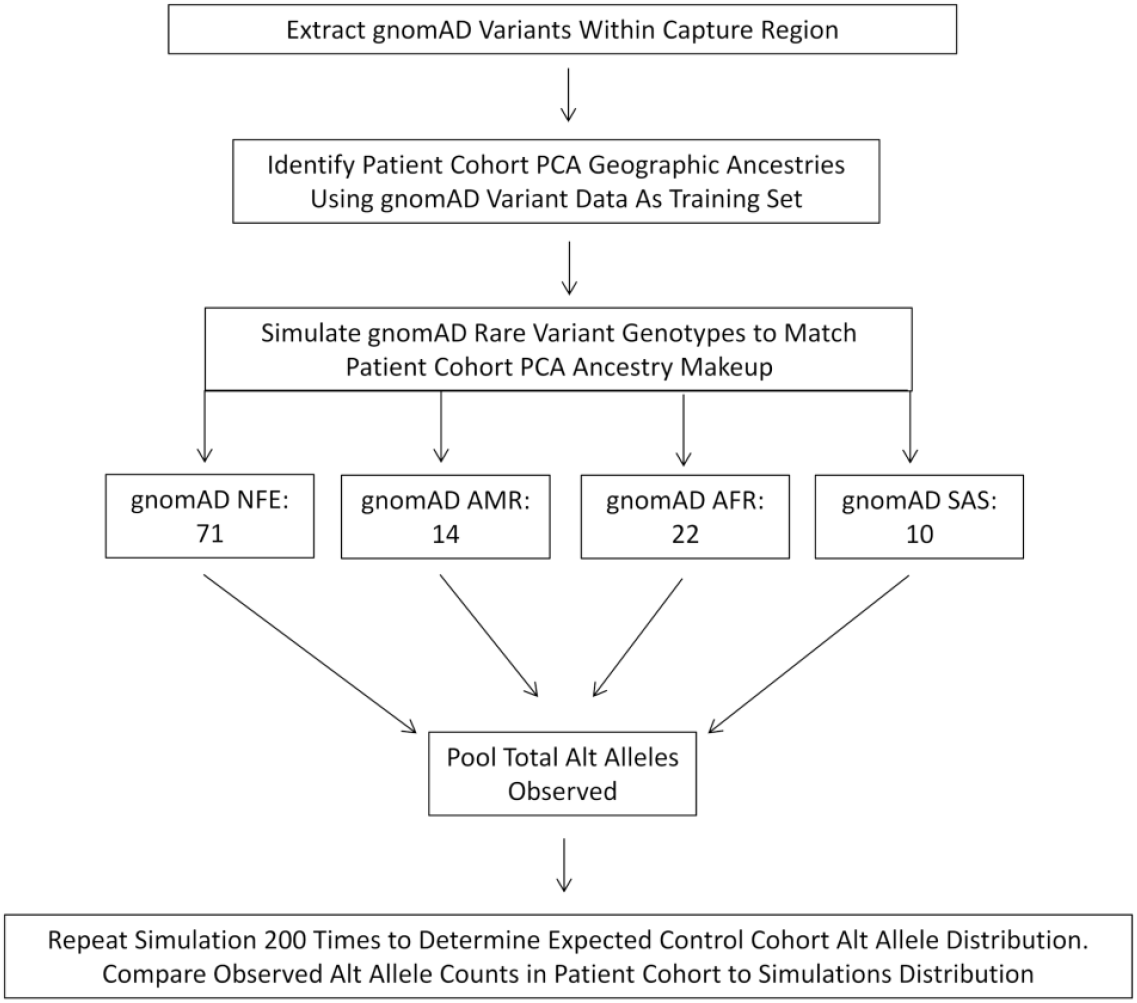
Experimental design for control simulation. Shown is a diagram of the experimental design for simulating controls. We first had to filter ExAC and gnomAD sites to only regions included in our targeted capture. Once the sites were filtered, we simulated training data to predict population ancestries for our cohort. We then simulated control rare variants for a control population with ancestries matched to our cohort, counting the total number of observed alternate alleles per gene. The process was repeated a total of 200 times to estimate the number of variants to expect on average for each gene.

### 2.4. Gamma-secretase & Notch pathway rare variants do not explain the majority of HS risk in our cohort

We first evaluated the observed set of protein-affecting variants for start-loss, premature stop codon, and frameshift variants predicted to be high impact loss-of-function changes. We observed two predicted loss-of-function variants, both in *NCSTN*, that to the best of our knowledge have not been reported in HS-affected individuals before: one frameshift variant (p.G6VfsTer22) and one premature termination variant (p.Q418*). A nearby premature termination variant, p.Q420*, has previously been reported to segregate with HS in a large autosomal-dominant kindred^30^. The American College of Medical Genetics and the Association for Molecular Pathology have provided guidelines for the interpretation of genetic variation with regards to likely pathogenicity^31^. Both of these *nicastrin* variants would be considered PVS1 (very strong) evidence of pathogenicity since this is a known HS mechanism and the variants aren’t at the far 3’ end of the protein (at ∼59% of protein length). These variants also alter protein length (PM4 moderate evidence). Large-scale population frequency data supports that loss-of-function variants are not tolerated in *nicastrin*. The gnomAD online database (controls v2.1.1) calculated that *nicastrin* has a 99.9% chance of being loss-of-function intolerant. Given the published papers demonstrating loss-of-function in *nicastrin* in HS, the evidence of pathogenicity for our cohort’s variants, and the likelihood of *nicastrin* being intolerant to these types of changes, we believe that the LoF variants in our cohort have a functional impact.

There were a greater number of rare synonymous or missense variants than loss-of-function variants in our cohort. However, the effects of synonymous and missense changes are more difficult to interpret, so we tested for an increased burden in each gene using control simulation. First, we tested genes for an increased burden of synonymous variants (**Fig 3**; **Table ST9**). We couldn’t exclude testing synonymous variants, as they can reduce protein half-life by introducing non-optimal codons, disrupt DNA binding motifs, or act as splice enhancers. No gene in our cohort had significantly increased synonymous variation.

**Figure 3.**
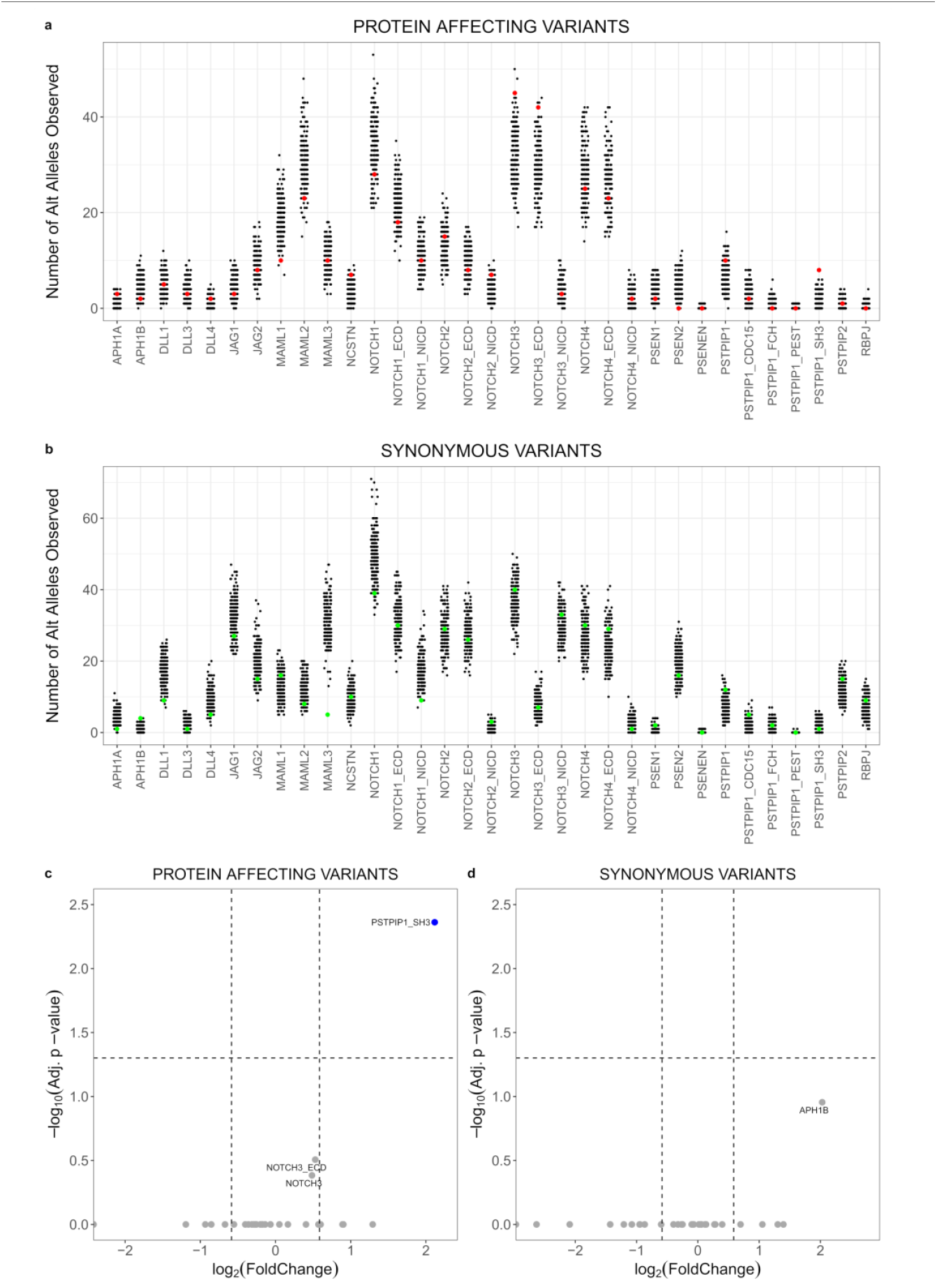
Variant enrichment burden test visualization. **a**. The simulation results for the expected number of rare variants per gene that alter amino acids. Each black dot represents the number of total minor alleles in a single simulation. The actual number of rare variants observed in our cohort is overlaid as a red dot. **b**. This panel has the same layout but is for synonymous variant burden, and our cohort observations are in green. **c**. The x-axis shows missense and synonymous coding variants (one dot per gene). The y-axis shows how many standard deviations from the mean our observations are compared to the control simulation. The majority are clustered around the simulation mean. **d**. The x-axis shows the variants as either missense or synonymous, and the y-axis is the adjusted P-value. The dashed line is a corrected P-value of 0.05. One test is highlighted, showing the PSTPIP1 SH3 domain which passed corrected significance.

Protein-affecting variants are perhaps easier to understand as they can alter protein structure and function. There was not a significantly increased burden for any of the gamma-secretase genes (**Fig. 3**; **Table 2**). Both *APH1A* and *NCSTN* had nominally significant increases in the number of missense variants. The three *APH1A* variants were concentrated at the c-terminus of the protein and predicted to be in either cytoplasmic domains or at the cytoplasmic end of a transmembrane domain^32^. One of these rare variants, p.C245W, was previously observed in an HS patient from another study and it is rare in non-Finnish Europeans^33^. There was also nominal significance for the *NOTCH2* intracellular domain, *NOTCH3*, and the *NOTCH3* extracellular domain. None of the Notch and Notch-related genes reached corrected statistical significance. Further study would be required to determine if any of these nominal enrichments is suggestive of a real increased burden. Regardless, rare missense variants in gamma-secretase and Notch-related genes do not explain much of the genetic risk for HS in our cohort.

**Table 2.** Rare missense variant burden analysis. Rare variant analysis results for capture panel genes and specific protein domains. The observed column lists the number of rare missense variants observed for the HS cohort. The simulated mean and SD values list the mean and standard deviation of the number of observed rare missense variants from 200 permutations of the synthetic control cohort. Raw and adjusted P-value columns show the uncorrected and Bonferroni corrected P-values, respectively, from the Poisson test. Several genes show raw significance, but only the *PSTPIP1* SH3 domain has a significant corrected P-value. Abbreviations: SD, standard deviation; Adj, adjusted; ECD, extracellular domain; NICD, Notch intracellular domain.

| Gene | Observed | Mean simulated | SD simulated | Raw P-value | Adj P-value |
| --- | --- | --- | --- | --- | --- |
| APH1A | 3 | 1.24 | 1.11 | 3.69E-02 | 1.00E+00 |
| APH1B | 2 | 4.64 | 2.15 | 1.00E+00 | 1.00E+00 |
| DLL1 | 5 | 4.65 | 2.16 | 3.22E-01 | 1.00E+00 |
| DLL3 | 3 | 3.60 | 1.90 | 1.00E+00 | 1.00E+00 |
| DLL4 | 2 | 1.31 | 1.14 | 1.44E-01 | 1.00E+00 |
| JAG1 | 3 | 4.50 | 2.12 | 1.00E+00 | 1.00E+00 |
| JAG2 | 8 | 9.64 | 3.10 | 1.00E+00 | 1.00E+00 |
| MAML1 | 10 | 18.97 | 4.35 | 1.00E+00 | 1.00E+00 |
| MAML2 | 23 | 30.63 | 5.53 | 1.00E+00 | 1.00E+00 |
| MAML3 | 10 | 10.96 | 3.31 | 1.00E+00 | 1.00E+00 |
| NCSTN | 7 | 3.94 | 1.98 | 4.74E-02 | 1.00E+00 |
| NOTCH1 | 28 | 33.69 | 5.80 | 1.00E+00 | 1.00E+00 |
| NOTCH1 ECD | 18 | 22.70 | 4.76 | 1.00E+00 | 1.00E+00 |
| NOTCH1 NICD | 10 | 11.53 | 3.40 | 1.00E+00 | 1.00E+00 |
| NOTCH2 | 15 | 13.57 | 3.68 | 2.88E-01 | 1.00E+00 |
| NOTCH2 ECD | 8 | 9.94 | 3.15 | 1.00E+00 | 1.00E+00 |
| NOTCH2 NICD | 7 | 3.67 | 1.91 | 3.36E-02 | 1.00E+00 |
| NOTCH3 | 45 | 33.43 | 5.78 | 2.24E-02 | 7.40E-01 |
| NOTCH3 ECD | 42 | 29.45 | 5.43 | 1.12E-02 | 3.71E-01 |
| NOTCH3 NICD | 3 | 3.38 | 1.84 | 1.00E+00 | 1.00E+00 |
| NOTCH4 | 25 | 28.14 | 5.30 | 1.00E+00 | 1.00E+00 |
| NOTCH4 ECD | 23 | 25.66 | 5.07 | 1.00E+00 | 1.00E+00 |
| NOTCH4 NICD | 2 | 1.85 | 1.36 | 2.83E-01 | 1.00E+00 |
| PSEN1 | 2 | 3.70 | 1.92 | 1.00E+00 | 1.00E+00 |
| PSEN2 | 0 | 4.93 | 2.22 | 1.00E+00 | 1.00E+00 |
| PSENEN | 0 | 0.07 | 0.25 | 1.00E+00 | 1.00E+00 |
| PSTPIP1 | 10 | 6.23 | 2.49 | 5.26E-02 | 1.00E+00 |
| PSTPIP1 CDC15 | 2 | 3.22 | 1.79 | 1.00E+00 | 1.00E+00 |
| PSTPIP1 FCH | 0 | 0.84 | 0.91 | 1.00E+00 | 1.00E+00 |
| PSTPIP1 PEST | 0 | 0.06 | 0.24 | 1.00E+00 | 1.00E+00 |
| PSTPIP1 SH3 | 8 | 1.85 | 1.36 | 1.32E-04 | 4.35E-03 |
| PSTPIP2 | 1 | 0.89 | 0.94 | 2.22E-01 | 1.00E+00 |
| RBPJ | 0 | 0.38 | 0.61 | 1.00E+00 | 1.00E+00 |

### 2.5. HS patients have an increased burden of rare variants in the SH3 domain of *PSTPIP1*

PSTPIP1 and PSTPIP2 belong to the F-BAR family of proteins, containing Fer/Cip4 homology (**FCH**) domains and BAR domains at their N-terminus. They can bind directly to lipid membranes through their F-BAR domains and serve as scaffolds for recruiting other proteins. Through their scaffolding functions, they are involved in the regulation of various membrane processes including endocytosis, phagocytosis, and both filopodium and podosome dynamics^34; 35^. Specifically, PSTPIP1 and PSTPIP2 have been shown to play a role in podosome dynamics in osteoclasts and monocytes^36; 37^. PSTPIP1 has also been shown to play a role in the regulation of T-cell activation^38; 39^. Variants in *PSTPIP1* are associated with the autoinflammatory diseases PAPA syndrome and familial recurrent arthritis in humans, while variants in *PSTPIP2* have been demonstrated to result in autoinflammatory disease in mice^23; 40; 41^. Since some individuals with PAPA-like syndromes also exhibit HS, we chose to evaluate *PSTPIP1/2* in our capture panel.

There was no evidence for protein-affecting variant enrichment in *PSTPIP2*. There were several rare missense variants in *PSTPIP1* that didn’t together meet significance for enrichment for the whole gene, but we noted that most of the variants were concentrated near the c-terminus of the protein. PSTPIP1 has four predominant protein domains: an FCH domain, a CDC15 domain, a PEST domain, and an SH3 domain, proceeding from n-terminus to c-terminus. We evaluated each of these regions independently under the hypothesis that distinct phenotypes could arise from variants affecting different domains. The FCH, CDC15, and PEST domains did not have an increased burden. The number of *PSTPIP1* SH3 domain variants observed in our HS cohort was significantly increased compared to the control simulations (Adjusted p-value = 4.35 × 10^−3^).

This enrichment resulted from four variants: p.T371I, p.A382T, p.G403E, and p.R405C (**Fig. 4**; **Table 3**). Seven individuals in our cohort (∼6% of the cohort) had one of these four variants, including one person homozygous for the A382T variant. To the best of our knowledge, T371I and A382T have not been previously reported in published case studies of either HS or PAPA-like syndromes. While the G403E variant has not been reported in these diseases before either, an individual with PAC (pyoderma gangrenosum, acne, ulcerative colitis) syndrome was previously reported to carry a *PSTPIP1* variant at the same amino acid position (p.G403R)^42^. The last SH3 domain variant that we observed, R405C, has been previously reported in two patients with some PAPA syndrome symptoms. One individual had pyoderma gangrenosum, acne conglobota, and suppurative hidradenitis (**PASH**), while the other individual was only reported to have pyoderma gangrenosum^37; 43^. Many of the known PAP syndrome variants, such as p.A230T and E250Q, are in the CDC15 domain. The observation of SH3 domain variants in patients with phenotypes similar to, but distinct from, PAPA syndrome suggests that there may be domain-specific phenotypes.

**Figure 4.**
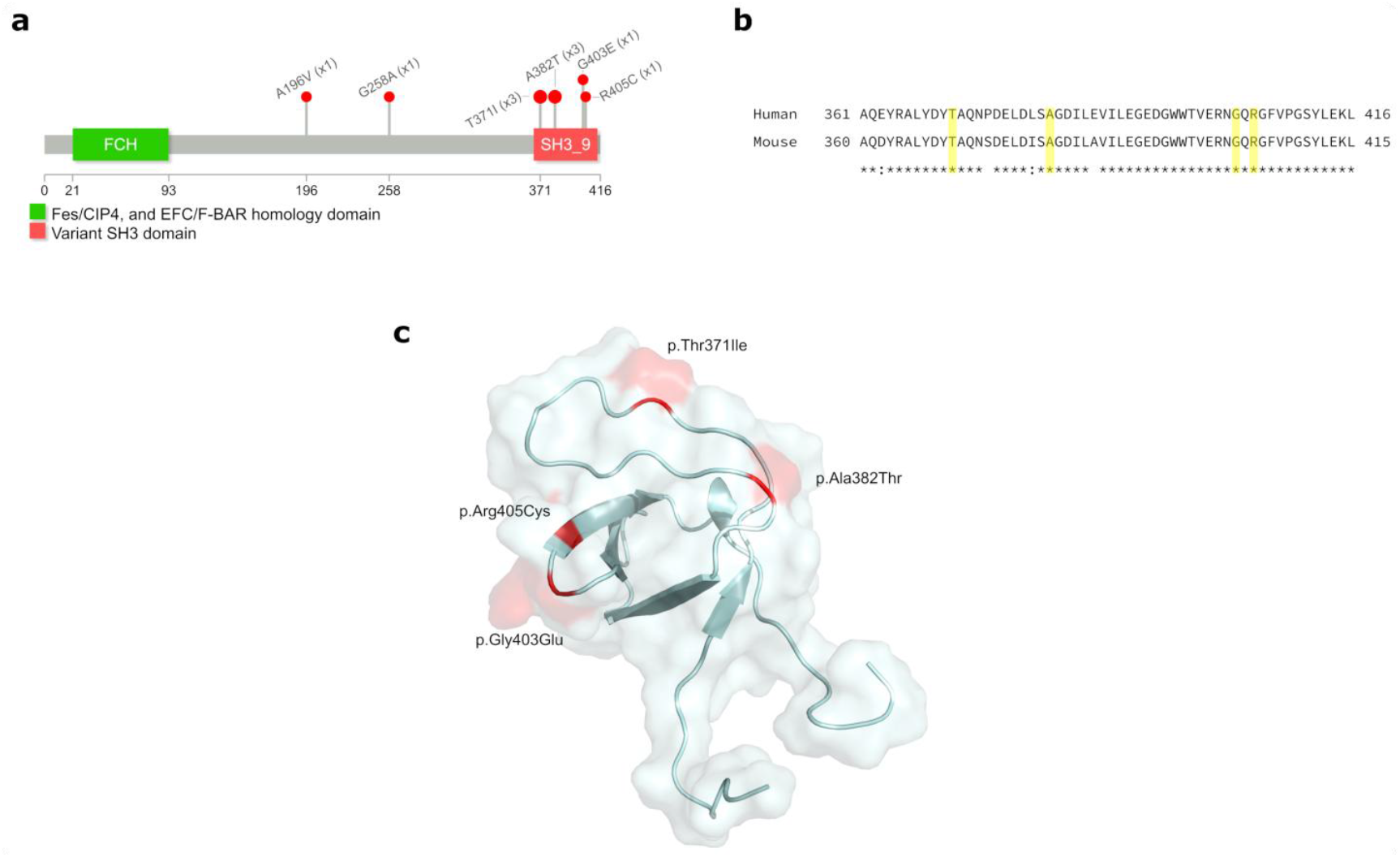
Localization of PSTPIP1 rare variants. **a**. A lollipops plot of the PSTPIP1 protein. While two variants are near the mid-point of the protein, the majority of identified variants are in the SH3 domain. **b**. Pairwise alignment of the amino acid sequences of PSTPIP1 in mice and humans shows the terminal portion of the protein shares a high identity. All four missense variants are in amino acids that are conserved in mice. **c**. The SH3 domain crystal structure shows the position of the missense variants in three dimensions. The folding of the protein puts the variants in closer physical proximity than one might expect from the linear sequence. They also appear to be surface-exposed and could alter the binding of this domain to different targets.

**Table 3.** *PSTPIP1* rare variants identified in our cohort. Shown are the variants in *PSTPIP1* with chromosome, position, reference / alternate alleles, cDNA and amino acid changes, number of allele observations, and number of individuals with at least one allele. The AFR, AMR, EAS, NFE, and SAS columns are the gnomAD allele frequencies of the variants. Abbreviations: chrom, chromosome; ref, reference allele; alt, alternate allele; AFR, African / African American population; AMR, Admixed American / Latino; EAS, East Asian; NFE, Non-Finnish European; SAS, South Asian.

| Chrom | Position | Ref | Alt | Domain | cDNA | Amino Acid | Alleles | Individuals | AFR | AMR | EAS | NFE | SAS |
| --- | --- | --- | --- | --- | --- | --- | --- | --- | --- | --- | --- | --- | --- |
| 15 | 77,322,867 | C | T | CDC15 | c.587C>T | p.Ala196Val | 1 | 1 | 6.26E-04 | 2.56E-04 | 5.13E-05 | 1.58E-05 | 2.62E-04 |
| 15 | 77,324,670 | G | C | CDC15 | c.773G>C | p.Gly258Ala | 1 | 1 | 3.29E-02 | 1.51E-02 | 2.06E-02 | 1.98E-03 | 2.81E-03 |
| 15 | 77,328,269 | C | T | SH3 | c.1112C>T | p.Thr371Ile | 3 | 3 | 1.17E-02 | 6.25E-04 | 0.00E+00 | 1.01E-04 | 6.93E-05 |
| 15 | 77,329,410 | G | A | SH3 | c.1144G>A | p.Ala382Thr | 3 | 2 | 1.47E-02 | 4.53E-04 | 5.12E-05 | 3.96E-05 | 3.27E-05 |
| 15 | 77,329,474 | G | A | SH3 | c.1208G>A | p.Gly403Glu | 1 | 1 | 8.35E-05 | 5.67E-05 | 0.00E+00 | 4.10E-04 | 0.00E+00 |
| 15 | 77,329,479 | C | T | SH3 | c.1213C>T | p.Arg405Cys | 1 | 1 | 4.61E-04 | 1.28E-03 | 0.00E+00 | 6.40E-04 | 3.28E-04 |

### 2.6. The SH3 domain variants observed in our cohort do not disrupt common PSTPIP1 binding partners

In macrophages, knock-down of *PSTPIP1* expression results in an increased number of podosomes^37^. In the same study, overexpression of wildtype *PSTPIP1* rescued podosome number, but overexpression of the R405C variant eliminated podosomes in favor of increased filopodial formation. This effect of the R405C variant was suggested to be a result of misregulation of WASp, as the authors showed that the R405C variant reduced binding of PSTPIP1 to WASp. Based on this previous data regarding the R405C variant, we wanted to test whether the PSTPIP1-SH3 domain variants we observed in our HS cohort affect the binding of PSTPIP1 to WASp. We performed co-immunoprecipitations by first exogenously expressing epitope (FLAG) tagged wildtype and variant PSTPIP1 in HEK293 cells and immunoprecipitating the PSTPIP1 protein. In addition to testing the constructs carrying the PSTPIP1 variants we detected in our HS cohort, we also evaluated a PSTPIP1 expression construct with the SH3 domain deleted as a control for complete loss of SH3 interactions. We then incubated the immunoprecipitated PSTPIP1 protein with cell lysates generated from THP-1 cells, a monocytic leukemia cell line. Co-immunoprecipitated proteins were analyzed by western blot (**Fig 5**). As previously reported, we found that wildtype PSTPIP1 bound to WASp. This interaction was dependent on the SH3 domain, as the SH3 domain deletion failed to precipitate WASp. Also consistent with previous results, we observed that the R405C variant reduced the binding of PSTPIP1 to WASp. The other HS variants didn’t have a clear trend. T371I had binding similar to wildtype. Both A382T and G403E appeared to have increased binding, though it’s worth noting that western blots aren’t quantitative. It does, however, support that the other HS variants don’t disrupt WASp binding as R405C does.

**Figure 5.**
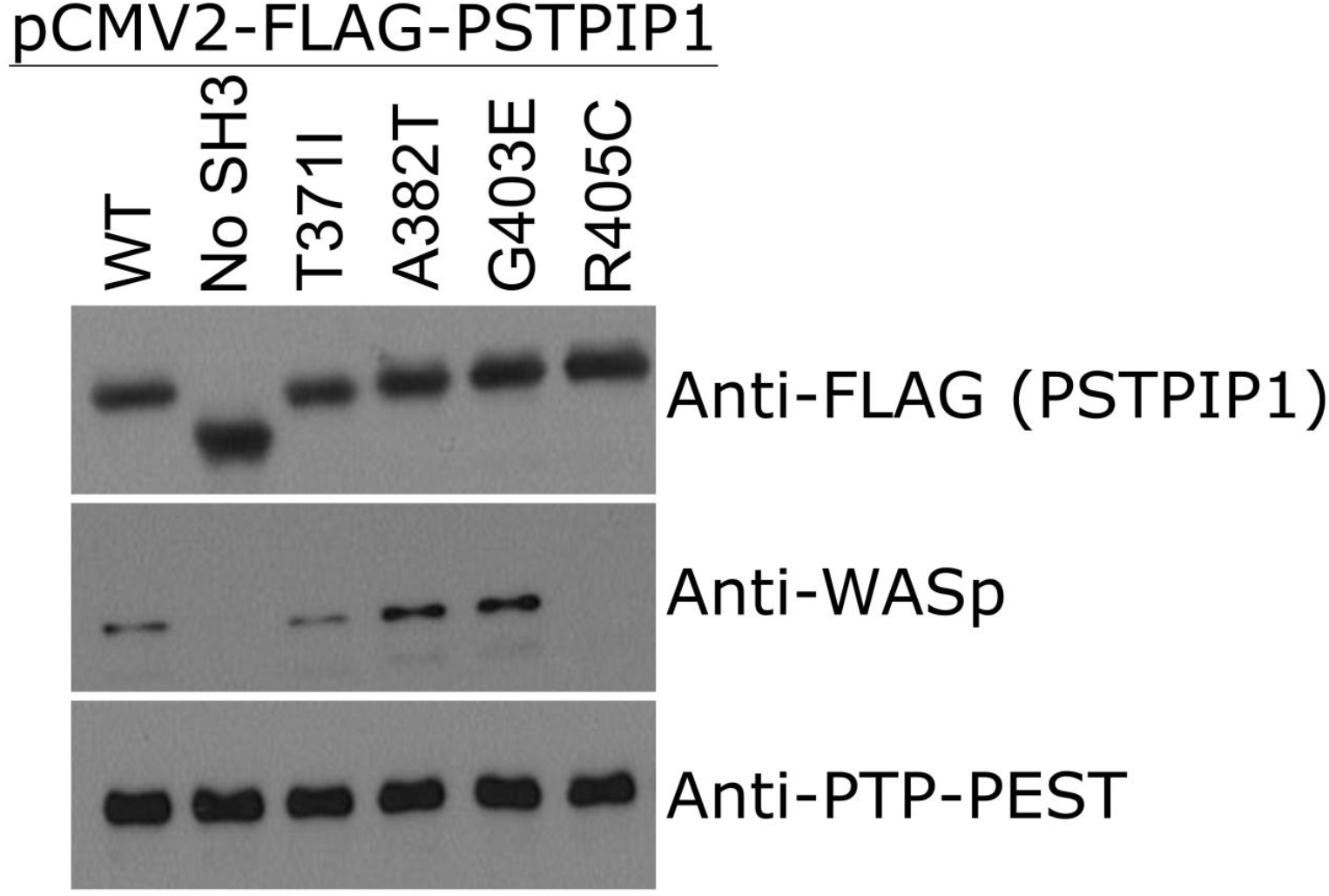
Western blots of PSTPIP1 wildtype and variant interactions with known binding partners. Western blot of binding partners immunoprecipitated by FLAG-tagged PSTPIP1. The variants tested included wildtype (**WT**) PSTPIP1 and construct missing the entire SH3 domain (No SH3) as a negative control for SH3 domain interactions. The other 4 SH3 variants (T371I, A382T, G403E, and R405C) were detected in our HS cohort and generated with site-directed mutagenesis. The anti-FLAG blot (upper panel) shows that the PSTPIP1 protein was expressed and that the SH3 domain deletion migrated faster since it was smaller. The anti-WASp blot (middle panel) showed intact binding between PSTPIP1 and WASp in wildtype, A382T, and G403E variants. The A382T and G403E variants may have slightly enhanced binding. The SH3 domain deletion and R405C failed to bind to WASp. The anti-PTP-PEST blot (lower panel) confirmed that the interaction between PTP-PEST and PSTPIP1 is SH3 domain independent. All PSTPIP1 variants, including the SH3 domain deletion, are bound normally to PTP-PEST.

We also evaluated the binding of these variants to the protein tyrosine phosphatase PTP-PEST, a protein known to interact with PSTPIP1^23^. Our data supported that PSTPIP1 binds to and can pull down PTP-PEST. The construct without the SH3 domain pulled down PTP-PEST also, supporting that the interaction is not dependent on that domain. None of the SH3 domain variants in our HS cohort disrupted PTP-PEST binding. Overall these data suggest that the newly reported HS variants may act through a distinct mechanism.

### 2.7 Deletions are not a common source of loss-of-function in gamma-secretase genes

The association of HS with loss-of-function in gamma-secretase genes is predicted from the observation of frameshift variants, early stop variants, and splice-affecting variants in individuals with familial HS. Loss-of-function can also occur through larger deletions. The role of genomic structural variation as it relates to gamma-secretase loss-of-function in HS is not well-characterized. To our knowledge, only one study has attempted to specifically evaluate structural variation in an HS cohort. An analysis of 48 HS-affected individuals by a multiplex ligation phosphorylation assay failed to find deletions in *NCSTN, PSENEN*, or *PSEN1*^21^.

Differential sequencing coverage between samples in a targeted capture is affected by capture batch and sequencing batch variability. The multiplexing of samples with the MDiGS capture method reduces this variability by capturing and sequencing multiple samples in the same batch. As demonstrated in the original publication describing the MDiGS method, this allows for sequencing coverage between samples to be normalized and compared to evaluate relative copy number variation (**CNV**). We took advantage of this feature of mDiGS to investigate whether we could identify any large deletion or insertion events in our HS cohort. Our process for identifying CNVs is described in detail in the Methods section, and each capture batch was analyzed separately. Briefly, we smoothed the per-base coverage in a 50 bp sliding window, calculated a coverage ratio by dividing the smoothed coverage by the average coverage for the batch, and segmented it into discrete copy number regions.

We evaluated the effectiveness of our CNV calling pipeline for identifying polymorphic CNVs in the gnomAD structural variant database that would be likely to be found in our cohort. We defined a CNV as polymorphic if the alternate allele frequency was at least 1% in all available ancestry sub-populations. We also added a length threshold, requiring structural variations to be at least 500 bp. Four polymorphic CNVs within our capture region met these criteria. We were able to identify instances of 3 of the 4 CNVs within our cohort (**Table 4; Fig. S4-S6**). We observed no evidence for the fourth polymorphic CNV, a 719 bp deletion in the *NOTCH2* region. There may have been no instances of this CNV in our cohort, though it is also possible that the smaller size of this deletion made it harder to detect using the comparative coverage method. The CNVs located in the *RBPJ* and *MAML2* capture regions were detected in our cohort at frequencies similar to the alternate allele frequencies reported in the gnomAD database. Interestingly, we observed the *PSTPIP1* deletion at a higher alternate allele frequency as is reported in the gnomAD database. Importantly, none of these polymorphic CNVs overlap exons for our genes of interest. This fact coupled with their relatively common minor allele frequency suggests that they are not strongly deleterious.

**Table 4.** Detected deletions and amplifications using relative coverage rates. Summary information about structural variants detected in our HS cohort and overlapping variants from gnomAD. In each case, the type of variant, gene it overlaps (if any), and chromosome are listed. The chromosome start and end, as well as estimated total size, is listed for both this cohort and from the gnomAD structural variation database. For gnomAD, the allele frequencies are listed for each of the major subpopulations. Abbreviations: HS, hidradenitis suppurativa; chrom, chromosome; alt freq, alternative allele frequency; AFR, African / African American population; AMR, Admixed American / Latino; EAS, East Asian; NFE, Non-Finnish European.

| Type | Gene | Chrom | HS Targeted Capture |  |  |  | gnomAD |  |  |  |  |  |  |
| --- | --- | --- | --- | --- | --- | --- | --- | --- | --- | --- | --- | --- | --- |
|  |  |  | Start | End | Size | Alt Freq | Start | End | Size | AFR | AMR | EAS | NFE |
| Amplification | NOTCH2 | 1 | 120,541,671 | 120,554,471 | 12,800 | 4.27E-03 |  |  |  | 0.00E+00 | 0.00E+00 | 0.00E+00 | 0.00E+00 |
| Deletion | Intergenic | 1 |  |  |  | NA | 120,675,954 | 120,676,673 | 719 | 1.93E-01 | 6.12E-01 | 4.45E-01 | 6.46E-01 |
| Deletion | RBPJ | 4 | 26,255,112 | 26,256,862 | 1,750 | 4.27E-03 | 26,255,264 | 26,256,575 | 1,311 | 1.50E-01 | 1.35E-02 | 0.00E+00 | 5.51E-03 |
| Deletion | RBPJ | 4 | 26,289,262 | 26,295,512 | 6,250 | 5.13E-02 | 26,290,469 | 26,295,385 | 4,916 | 4.94E-02 | 3.63E-02 | 9.77E-02 | 5.23E-02 |
| Deletion | MAML2 | 11 | 96,001,925 | 96,003,475 | 1,550 | 1.54E-01 | 96,001,925 | 96,003,394 | 1,469 | 2.22E-01 | 3.07E-01 | 1.54E-01 | 1.04E-01 |
| Deletion | PSTPIP1 | 15 | 77,330,643 | 77,332,793 | 2,150 | 5.26E-01 | 77,330,745 | 77,332,721 | 1,976 | 1.34E-01 | 3.67E-01 | 2.16E-01 | 4.22E-01 |

We detected two rare CNVs in our capture cohort: one in the *RBPJ* capture region and one in the *NOTCH2* capture region. The *RBPJ* capture region deletion is upstream of the first exon (**Fig. S7**). The *NOTCH2* region CNV is an amplification that spans an exon of *NOTCH2* (**Fig. S8**). It’s unclear what effect this amplification would have on the final protein, and if the protein would be stable. Both of these CNVs were observed in only one individual each in the cohort. Overall, we did not detect any substantial prevalence of exon-overlapping deletions or amplifications for genes in our capture panel.

## 3. Discussion

Human phenotypes are complex and influenced by environmental exposures, relatively small effects from common polymorphic variants, potentially large effects from rare genetic variants, and the interplay between all of those factors. For hidradenitis, known environmental risk factors include smoking and obesity. Currently, there are no well-powered genome-wide association studies to identify common variants linked to risk for HS. Rare variants in genes involved in the gamma-secretase complex are known, high-penetrance risk factors for autosomal dominant HS. We discovered two new nicastrin loss-of-function variants in this cohort, but only in two individuals. This is consistent with a study looking at genetic variants in *NCSTN* in 95 HS patients, where only one premature termination variant was observed^22^. Another possibility for gamma-secretase, especially nicastrin, loss-of-function is larger-scale structural variation, i.e. only some individuals have frameshifts, canonical splice site alteration, start-loss, and premature stops, but more have a deletion of multiple exons or the entire gene. In our cohort, deletions were not a major contributor to loss-of-function in gamma-secretase. This finding, importantly, decreases the likelihood that many individuals with HS carry gamma-secretase gene deletions. A final way that gamma-secretase could contribute a greater risk to sporadic HS is through an increased burden of loss-of-function via rare missense variants. We failed to find a statistically significant accumulation of rare protein-affecting variants in any gamma-secretase gene, though both *APH1A* and *NCSTN* were nominally significant. Taken together these data support two findings in the published literature: 1) loss-of-function in nicastrin is a high-penetrance risk factor for HS, but 2) most individuals with HS do not have loss-of-function in gamma-secretase genes.

A popular theory is that the loss-of-function in gamma-secretase causes HS via altered Notch signaling that disrupts skin development and homeostasis. We did not find damaging loss-of-function variants, excess structural variation, or a statistically significant accumulation of rare protein-affecting variants in any of our captured Notch-related genes. This doesn’t exclude a role for Notch signaling in HS but does suggest that if it has a role it’s more subtle than genetic haploinsufficiency of a single Notch signaling gene. This is perhaps not unexpected since Notch signaling plays important and diverse roles during embryonic development and across the lifespan. For example, rare genetic variation, including loss-of-function, of *NOTCH1* is associated with Adams-Oliver syndrome 5 (OMIM # 616028) and aortic valve disease 1 (OMIM #109730). Both of these genetic syndromes are arguably more severe (and in the case of Adams-Oliver syndrome exhibit greater phenotypic pleiotropy across tissues) than what is observed in HS. Our findings don’t exclude a role for Notch signaling in HS, but support the possibility that gamma-secretase haploinsufficiency may operate through more diverse mechanisms.

There are multiple lines of evidence, such as clinical response to anti-TNF biologics, overlap with other inflammatory diseases, and higher prevalence in women (consistent with other autoimmune/inflammatory diseases), which support a primary role for enhanced inflammation in HS^1-3; 54; 55^. This is particularly intriguing with our finding of an increased burden of rare protein-affecting variants of *PSTPIP1*. One of the first associations of *PSTPIP1* variants with human disease was in a study involving two families, one with PAPA syndrome and the other with familial recurrent arthritis^23^. Since then, numerous case reports have described *PSTPIP1* rare variants in people with symptoms consistent with PAPA syndrome, as well as various PAPA-like syndromes^42-50^. These PAPA-like syndromes include cases with overlap between PAPA syndrome phenotypes and HS. One example is PASH syndrome, with pyoderma gangrenosum, acne conglobota, supportive hidradenitis, but no arthritis. Another example is PAPASH syndrome, which includes all the symptoms of PAPA syndrome along with HS^25^. It’s unclear clinically whether these individuals have overlapping phenotypes, i.e. both HS and PAPA syndrome co-existing as separate entities, or if HS and PAPA syndrome is part of a single inflammatory disease spectrum. Our HS cohort was specifically enriched for variants in the SH3 domain of PSTPIP1. This hints at the possibility that function-altering variants in *PSTPIP1* might form an allelic series with PAPA syndrome at one extreme and isolated features, such as HS or acne conglobota, at the other depending on the specific impact of individual variants.

It is important to note while all three cohorts were ascertained specifically for HS, individuals were not specifically included or excluded because of other inflammatory features. For the SVUH and WU cohorts, we don’t have additional clinical information. Two of the individuals with SH3 domain variants were in the OSU cohort, for which we were able to gain additional information. One OSU cohort individual had the T371I variant. Review of their clinical history noted a history of severe acne and joint pain, though they were only diagnosed with hidradenitis. The other OSU individual with an SH3 domain variant had R405C. This variant has been previously observed in a patient with aggressive pyoderma gangrenosum and in another patient with PASH syndrome^37; 43^. A review of our patient’s history showed chronic HS, but no evidence for arthritis, acne conglobota, or pyoderma gangrenosum. It’s unlikely that many individuals with SH3 variants in our cohort also have full-blown PAPA syndrome, as PAPA syndrome is an orphan disease with few patients identified worldwide. Also of interest, there is a report of an individual with PASH who had a loss-of-function variant in *NCSTN*^51^. These findings further support both phenotypic and genetic overlap between HS and PAPA syndrome phenotypes.

If different types of missense variants in *PSTPIP1* produce an allelic series, the next logical question is what effect our identified SH3 variants have. We found R405C disrupted binding to WASp, consistent with published literature. None of the other HS variants disrupted WASp binding, and none of the variants disrupted binding to PTP-PEST. This may not be unexpected, as the HS-associated variants may act by different mechanisms. Another approach to this question is examining the existing literature for these types of variants. The *PSTPIP1* R405C variant that we observed has been reported in two individuals with PAPA-like disease. In one study, the patient had symptoms consistent with PASH, thus providing an example of this variant occurring in an individual with HS^43^. In another study, a patient with R405C presented only with pyoderma gangrenosum, though it was noted that the patient’s father had a history of severe acne^37^. The authors were able to show that this variant abrogated the ability of PSTPIP1 to bind to WASp, resulting in dysregulation of podosome formation in macrophages. Although the *PSTPIP1* G403E variant that we observed in our cohort has not been reported in any patients with PAPA-like disease, a patient with a p.G403R variant was reported as presenting with a non-classical PAPA-like syndrome, exhibiting pyoderma gangrenosum, acne, and ulcerative colitis^42^. These previously published reports lend some additional support to the idea that a range of different phenotypes affecting multiple tissues might be produced from different PSTPIP1 variants, and that some of these SH3 domain variants may substantially increase the risk of developing HS.

The PSTPIP1 SH3 domain has been shown in some functional studies to inhibit T-cell activation. One study observed that PSTPIP1 inhibits cell surface expression of CD69 and production of both IL-2 and interferon gamma after CD2 stimulation, but not after T-cell receptor stimulation^52^. This study found that overexpression of PSTPIP1 lacking the SH3 domain does not inhibit T-cell activation. Another study demonstrated that expression of PSTPIP1 in T-cells inhibits CD3-stimulated T-cell activation, but the expression of a p.W396A variant that abolishes many SH3 domain interactions resulted in increased T-cell activation^38^. A third study has shown that PSTPIP1 localizes to T-cell immunological synapses and that some PSTPIP1 variant constructs reduced synapse formation^39^. One possible link between these functions and HS is an alteration of T-cell activation. Previous work has demonstrated that one of the most common pathophysiological findings in early HS lesions is the presence of follicular plugs and a dense T-cell infiltrate^53^. Follicular plugging could lead to rupture of follicles and leaking of contents into the dermis. This might trigger a local T-cell response, which could be exaggerated by dysfunction in PSTPIP1-mediated inhibition. Further work is needed to identify PSTPIP1 binding partners in different cell types and to define the function of these rare SH3 domain variants in relevant cells.

Consistent with other cohorts, we can confirm that a small, but appreciable, percent of individuals with HS have a *NCSTN* loss-of-function rare variant (<2% in our cohort). Although we have identified an association with a new gene of interest, only ∼6% of our cohort carried a *PSTPIP1* rare missense variant in the SH3 domain. This suggests that much of the genetic risk for HS is unidentified. With limited resources, we had to develop a panel of candidate genes and work with a small cohort. Candidate gene screens are always biased in a way that whole-genome screens are not. Additional discovery will require genome-wide studies (rare variant sequencing and genome-wide SNP association) in larger HS cohorts. Our study has, however, reinforced that genetic variants can be important risk factors for the development of HS, and further supported inflammatory mechanisms in HS. Screens of larger cohorts may provide more resolution to determine if the genes that had only nominally significant variant enrichments in our study – *APH1A*, the *NOTCH2* intracellular domain, and the *NOTCH3* extracellular domain – may be real signals. Critically, for the HS community, there are few effective treatment options available. The delineation of genetic risk factors, their interplay with environmental triggers, and a better understanding of their roles in the molecular evolution of HS lesions are required to identify rational new treatment targets.

## 4. Methods

### 4.1. Genomic DNA isolation

#### Whole blood DNA isolation

We mixed 2 mL of blood with 2 mL of red blood cell lysis buffer (0.32 M Sucrose, 10.0 mM Tris-HCl, 5.0 mM MgCl_2_, 0.75% Triton X-100, pH 7.4), and 2 mL of ice-cold water, and mixed by inverting 6-8 times. We incubated the mix at room temperature for 3-5 minutes, followed by centrifugation at 2600 rcf for 15 minutes at 4°C. We discarded the supernatant and washed the pellet twice. Washes were performed by adding 2 mL of red blood cell lysis buffer and 6 mL of cold water to the pellet, mixing by vortexing, centrifuging at 2600 rcf for 15 minutes at 4°C, and discarding the supernatant. After two washes, we added 5 mL of proteinase K buffer (20 mM Tris-HCl, 4.0 mM EDTA-Na_2_, 100 mM NaCl, pH 7.4) and 0.5 mL of 10% SDS to the pellet and resuspended by vortexing. We added 50 µL of proteinase K (20 mg/mL) and mixed it by inverting. The proteinase K reaction was incubated at 55°C overnight. We then added 4.0 mL of 5.0 M NaCl and mixed by vortexing. The solution was centrifuged at 3400 rcf for 30 minutes at 4°C, and the supernatant was transferred to a new tube. We added an equal volume of ice-cold isopropanol to the supernatant, mixed by inverting several times, and then incubated on ice for 10 minutes. The mixture was centrifuged at 3400 rcf for 20 minutes at 4°C. We discarded the supernatant and carefully washed the pellet with 70% ethanol. After washing we carefully discarded the supernatant and air-dried the pellet for 5-10 minutes. Finally, we resuspended the pellet in 200-500 µL Qiagen Elution Buffer (Qiagen 19086).

#### Saliva DNA isolation

Saliva samples were collected from patients and preserved in Biometrica Salivagard containers until DNA isolation. We used Qiagen DNeasy Blood and Tissue Kits to isolate genomic DNA according to the manufacturer’s instructions.

### 4.2. DNA sequencing library creation

We sheared 600 ng of genomic DNA (10 ng/µL concentration) in 1X T4 DNA Ligase Buffer (Enzymatics L6030-HC-L) using a Covaris Sonicator (Duty Cycle 10%, Peak Incident Power 175, Cycles/Second 200, Time 85 seconds). We end-repaired the fragments by adding the following reagents to 50 µL of the sheared products: 2.25 µL T4 DNA Polymerase (Enzymatics P7080L), 2.25 µL T4 Polynucleotide Kinase (Enzymatics Y9040L), 1.0 µL 25 mM dNTP (Bioline BIO-39049), 0.55 µL 10X T4 Ligase Buffer Enzymatics L6030-HC-L). We incubated the reaction at 20°C for 40 minutes, and then at 75°C for 25 minutes to heat inactivate enzymes. We then added a dA-tail to the repaired fragments by adding the following to the heat-inactivated end-repair reaction: 0.65 µL 10 mM dATP (Enzymatics N2010-A-L), 0.5 µL 10X Taq B DNA Polymerase Buffer (Enzymatics P7250L), 1.9 µL Taq B DNA Polymerase (Ezymatics P7250L), and 1.95 µL molecular grade water. We incubated the dA-tailing reactions at 70°C for 25 minutes.

We ligated the dA-tailed fragments with Illumina TruSeq Y-adapters. We prepared the Y-adapters by incubating equimolar oligos at 95°C on a heat block for 5 minutes. We then turned off the heat block and allowed the Y-adapters to anneal as the mixture gradually cooled to room temperature. We added the following to the dA-tailed reaction: 18.5 µL 2X Rapid Ligase Buffer (Enzymatics L6030-HC-L), 1.5 µL 50 µM annealed adapters, 1.0 µL T4 Rapid DNA Ligase (Enzymatics L6030-HC-L), and 1.0 µL molecular grade water. We incubated the ligation reaction at 25°C for 20 minutes, then removed the excess unligated adapter from the reaction using 0.75X volumes of Agencourt AMPure XP beads. We washed beads twice with 80% ethanol and resuspended them in 32 µL Qiagen Elution Buffer to elute DNA. We then used 30 µL of the purified adapter-ligated fragments as templates for the indexing PCR reaction by adding the following: 11 µL 5 M Betaine (Sigma-Aldrich B0300-5VL), 11 µL 5X VeraSeq Buffer (Enzymatics P7511L), 0.5 µL 25 mM dNTP (Bioline BIO-39049), 1.0 µL VeraSeq 2.0 Polymerase (P7511L), 0.75 µL indexed i5 primer (100 µM), 0.75 µL indexed i7 primer (100 µM). All relevant sequences are listed in table **Supplementary Table ST7**. For the combinatorial indexing reaction, we denatured the DNA by incubating at 98°C for 30 seconds, followed by 14 cycles of 98°C denaturation for 10 seconds, 62°C annealing for 30 seconds, and 72°C extension for 45 seconds. We used a 5 minute final extension at 72°C. We added 20 µL Qiagen Elution Buffer to the reaction mixture, and then removed excess primer and nucleotides using 0.9X volumes of Agencourt AMPure XP beads. We washed beads twice with 80% ethanol and then resuspended them in 30 µL of Qiagen Elution Buffer to elute DNA.

### 4.3. Targeted genomic capture

We established a targeted capture protocol based on the MDiGS methodology with limited modification^27^. To generate capture probes, we acquired fosmids and bacterial artificial chromosomes (**BACs**) containing human DNA sequences spanning the genomic regions of interest. Fosmids/BACs were either purchased from the BACPAC Resource (Children’s Hospital Oakland Research Institute) or provided as a gift from Dr. Evan Eichler (University of Washington). The fosmid/BAC identifiers and the genomic regions they cover can be found in **Supplementary Table ST1**. We combined 50 – 75 ng of each fosmid/BAC into a single tube and then generated biotinylated capture probes using the Biotin-Nick Translation Mix (Roche Cat. No. 11745824910) following the manufacturer’s instructions. We then ran nick translation reactions over Roche G-50 Sepharose Quick Spin DNA Columns (Roche 11814419001), according to the manufacturer’s instructions, to remove unincorporated biotinylated nucleotides. We collected column elutions and used them as capture probes. Capture probe concentration was estimated as the nick translation input DNA divided by the sepharose column elution volume.

We mixed 200 ng of biotinylated fosmid/BAC probes with 5 µg of Human Cot-1 DNA (Invitrogen 15279011) and then lyophilized in a speed-vac. We resuspended the lyophilized probes in 3.0 µL of molecular-grade water and transferred the probes to a PCR tube. We incubated the probes at 95°C for 5 minutes and then at 65°C for 15 minutes. We then added 3.0 µL of 2X Hybridization Buffer (1.5 M NaCl, 40 mM sodium phosphate buffer [pH 7.2], 10 mM EDTA [pH 8.0], 10x Denhardt’s Solution [Invitrogen 750018], 0.2% SDS) to the probes, followed by incubation at 65°C for 5 hours.

We added 3.5 µL Post-PE1 primer (100 µM) and 3.5 µL Post-PE2 primer (100 µM) to 2.0 µg of pooled sequencing libraries, and lyophilized the libraries and primers in a speed-vac. In this step, Post-PE1 and Post-PE2 primers served to block DNA fragment chaining during capture hybridization. We resuspended the lyophilized sequencing libraries and primer mix in 3.0 µL water and incubated at 95°C for 5 minutes, followed by incubation at 65°C for 15 minutes. We added 3.0 µL 2X Hybridization buffer to the libraries and then mixed the libraries with the capture probes. We added a drop of oil (Sigma BioReagent Mineral Oil M59054) to cover the hybridization reaction and incubated it at 65°C for 80 hours. We pulled down the hybridized fragments using M-270 Streptavidin Dynabeads (Invitrogen 65305). We prepared the beads by washing them two times with Binding Buffer (10 mM Tris-HCL [pH 7.5], 1 mM EDTA [pH 8], and 1 M NaCl). After aspirating the Binding Buffer we resuspended the Dynabeads in 150 µL of fresh Binding Buffer, added the hybridization reaction to the Dynabeads, and incubated on a rocker at room temperature for 1 hour. We then washed the beads with 200 µL 1X with Wash Buffer 1 (1X Saline-Sodium Citrate Buffer [MilliporSigma S6639] with 0.1% SDS) on a rocker at room temperature for 15 minutes. We washed the beads three times with 200 µL Wash Buffer 2 (0.1X Saline-Sodium Citrate Buffer with 0.1% SDS) at 65°C for 5 minutes. We eluted the DNA from the beads by adding 50 µL of 0.1M NaOH for 10 minutes at room temperature. We transferred the supernatant to a new tube and neutralized the elution with 50 µL 1M Tris-HCl. We then purified DNA by binding to 180 µL of Agencourt AMPure XP beads. Beads were washed 2X with 80% ethanol and resuspended in 27 µL Qiagen Elution Buffer to elute DNA.

We amplified the post-capture sequencing libraries to get sufficient yield for sequencing. We used 25 µL of eluted capture DNA, 0.75 µL POST-PE1 (100 µM), 0.75 µL POST-PE2 (100 µM), 1.0 µL 10 mM dNTPs, 5X Q5 Polymerase Buffer, 0.5 µL Q5 Polymerase (New England Biolab M0493), 12.0 µL molecular-grade water. We initially denatured for 30 seconds at 98°C, followed by 13 cycles of denaturation at 98°C for 30 seconds, annealing at 60°C for 30 seconds, and extension at 72°C for 30 seconds. We used a 2 minute final extension at 72°C. We purified the products by adding 25 µL Qiagen Elution Buffer and 64 µL Agencourt AMPure XP beads. Beads were washed 2X with 80% ethanol and resuspended in 30 µL of Qiagen Elution Buffer.

### 4.4. Sequencing Analysis

Command details can be found in the supplement. We removed sequencing adapters and trimmed low-quality 3’ sequence with cutadapt v1.14^54^. We aligned the cleaned reads to the Broad GRCh37 version of the human genome that includes a decoy contig (human_g1k_v37_decoy.fasta) using bwa aln v0.7.17^55^. For genotyping, we processed the aligned reads using Genome Analysis Tool Kit (**GATK**) best practices with haplotype-based genotype calling^56; 57^. Due to the small size of our targeted capture regions, we used hard filters instead of Variant Quality Score Recalibration. Variants were filtered if they met any of the following conditions: QualByDepth < 2.0, ReadPosRankSum < -20.0, InbreedingCoeff < -0.8, FisherStrand>200.0, StrandOddsRatio>10.0.

### 4.5. Variant quality control

To generate data for a simulated control cohort, we utilized genetic data from gnomAD. We queried the gnomAD genomes and exomes sites files (v2.1.1) to collect all variants at genomic positions that fell within the regions covered by our fosmids. We then performed a reciprocal depth filter. We identified positions from our targeted capture and from the gnomAD variants where median coverage across all samples was less than 20. Positions below this depth cutoff in either the targeted capture or the gnomAD database were removed from both cohorts. Furthermore, positions where a genotype was not called for 10 percent or more of our HS patient alleles were removed from both the targeted capture analysis and the gnomAD variant tables. After filtering based on depth and unknown genotype, the gnomAD exome and genome variant tables were merged. gnomAD variants present in both the genome and exome sites files were compiled by summing alternate allele counts (**AC**) and total allele number (**AN**) for each ancestral population subgroup.

For our variant burden analysis, we only analyzed variants located in coding regions of our genes of interest. Annotations were performed for one selected transcript for each gene (**Supplementary Table ST2**). Transcripts were selected from Ensembl using a strategy that prioritized transcripts having a consensus coding sequence (**CCDS**) ID, followed by having a gold-labeled biotype categorization over red biotype, and finally by choosing the transcript that coded for the longest protein in cases where multiple transcript options remained. We used the Ensembl Variant Effect Predictor (VEP) to determine the predicted impact of the variants on amino acids. Protein-affecting and synonymous variants were retained for performing separate burden analyses. The code used to quality filtering variants, to collect coding region variants, and to run subsequent analyses is available at https://github.com/RobersonLab/2022_hs_pstpip1.

### 4.6. Classifying individual geographic ancestry by gnomAD simulation and principal components analysis

We leveraged subpopulation variant allele frequencies from gnomAD to classify our HS patient cohort into geographic ancestral populations using Principal Component Analysis (**PCA**). First, we simulated genotypes for individuals from various geographic ancestries using the following gnomAD variant allele frequencies: African/African American (**AFR**), Latino (**AMR**), East Asian (**EAS**), Non-Finnish European (**NFE**), and South Asian (**SAS**). Genetic variants were simulated using the ‘random’ package from the Python Standard Library (v2.7) as a random number generator. For each simulated variant, two random numbers between 0 and 1 were selected to represent an individual’s two alleles at the genomic position. Each random number was then challenged against the ancestry alternate allele frequency for the individual being simulated. If the random number was less than or equal to the challenge alternate allele frequency, the individual accrued an alternate allele at the position. If the random number was greater than the challenge alternate allele frequency the individual accrued a reference allele. For the PCA analysis, we limited simulated sites to variants that were identified in our HS patient cohort and were detected in the quality-filtered gnomAD exome variant set. We did not use the gnomAD genome variants because the SAS ancestry group was not provided in the gnomAD genome sites file due to the small sample size. We simulated all variants and did not limit our simulated sites to polymorphisms. We simulated genotypes for 2000 individuals per gnomAD subpopulation. We combined our cohort data with the simulated cohorts and converted the genotypes to an integer indicating 0, 1, or 2 “doses” of alternative alleles. We performed PCA on this matrix using ‘prcomp’ in R (v3.6.0). We also performed uniform manifold approximation and projection (**UMAP**) on the principal components 1-6 using the ‘umap’ package in R to plot the principal component data after dimension reduction. We assigned each individual in our cohort a gnomAD ancestry subpopulation using K-nearest neighbor (**k-NN**) clustering analysis. The simulated individuals were separated into two groups, both with 1000 individuals of each subpopulation: a “simulation” group and a “training” group. We used the first six principal components from these groups to train the ‘knn’ function in R. We tested the k-NN algorithm using the “simulation” group as the known truth set and the “training” group as the experimental “unknown” set. We evaluated k-NN ancestry assignment of the “training” group using different K values to determine the optimal K value that resulted in the fewest categorization errors. We determined the optimal K value for our analysis to be K=25. After K value optimization we then used k-NN to assign estimated gnomAD subpopulation ancestries to our HS affected cohort using the 1000 simulated individuals of each subpopulation in the “simulation” group as the training data.

### 4.7. Variant Burden Enrichment Testing

We simulated the expected number of rare variant genotypes for a control cohort using gnomAD data to determine if what we observed in our cohort might be statistically significant. The variant simulation was performed using random number challenge against alternate allele frequencies as described above; however, genotypes were simulated for rare synonymous or rare protein-affecting variants within the targeted capture region. A rare variant was defined as a variant that is observed at a minor allele frequency of less than or equal to 1.0% in at least one of the five gnomAD subpopulations evaluated (AFR, AMR, EAS, NFE, SAS). A variant that met this threshold for one population was included in the analyses for all populations to simulate the same set of variants for all populations. For multi-allelic genomic positions, all variants at the position that met our 1.0% alternate allele frequency threshold were combined to create a genomic position alternate allele frequency by summing all of the variant alternate allele frequencies. gnomAD variants for which only one alternate allele was observed amongst all five subpopulations were classified as singletons and were not simulated individually in the rare variant genotype simulations. Instead, singletons were used to calculate a gene-specific singleton rate for each gene by counting up the number of synonymous or missense singletons within a gene and dividing that number by the average total allele number that was sequenced in gnomAD at those singletons. Instead of simulating each singleton variant individually, for every simulated individual two random numbers, representing two alleles, were generated and challenged against the gene-specific singleton rate to determine if the individual would acquire a singleton alternate allele.

We simulated rare variant genotypes for 117 controls using gnomAD subpopulation alternate allele frequencies. The geographic ancestry assigned to the control cohort matched the same ratio as our HS cohort. We repeated this simulation 200 times to determine the distribution of the number of rare variants that would be expected in a cohort the size and makeup of our HS cohort. We used the mean number of rare variants observed for each gene or protein domain over the 200 simulations as the Poisson lambda parameter for statistical testing. We performed one-sided (greater than expectation) Poisson testing to determine the significance of the number of variants observed in our HS cohort compared to the simulations. We adjusted p-values by applying a Bonferroni correction for the total number of genes and protein domains that we evaluated.

### 4.8. PSTPIP1 visualizations

We generated the *PSTPIP1* gene lollipop plot using the lollipops program^58^. We determined the pair-wise alignment between human and mouse PSTPIP1 proteins using the Clustal Omega web tool (human uniprot O43586, mouse uniprot P97814). The 3D structure for the human SH3 domain was from the NCBI structure database, identifier 2DIL. We visualized this structure using open-source Pymol v1.8.4.0.

### 4.9. Copy Number Variation Analysis

We used the sequencing coverage of each sample to estimate whether they had any deletions or amplifications. Sequencing coverage within a sample varies over the length of a capture region as a result of total sequencing depth, local DNA context, and different capture efficiencies. We analyzed samples in batches based on when they were captured to account for capture efficiency differences. For each sample, we normalized each capture region by dividing it into 50 bp sliding windows and calculating average sequencing coverage over the window. Within 25 bp of the end of a capture region, the positions were pooled into the penultimate sliding window for the region. The sliding windows smoothed variation in sequencing coverage due to local DNA context. We determined a reference coverage value for each sliding window by averaging the smoothed coverage for all samples in the batch, assuming this average value was representative of 2 copies at the position. We calculated a coverage ratio by dividing a sample’s smoothed value for a given sliding window by the batch average coverage. We adjusted for unequal loading of the sample in the capture reaction by taking the sliding window coverage ratios and dividing them by the mean normalized ratio for all sliding windows within that sample across all capture regions. These final values were multiplied by 2 to convert them to the estimated copy number for the position.

The copy number estimates were segmented using the R ‘copynumber’ package (version 1.26.0) which identifies discrete segments across genomic regions and returns the average copy number for each individual in each segmentation. Each capture region was segmented independently. We normalized the average copy number for all individuals in all segmentations to the average copy number across all segmentations in the capture region being analyzed. We noted that some segmentation regions had polymorphic CNVs at a high enough frequency to skew the intra-segmentation mean copy number value away from the center of any CNV group, making it unclear which population, if any, was a CNV population. To account for this, we performed a second intra-segmentation normalization step in which we normalized the copy number for all individuals to a ‘modified median’ copy number within that segmentation. For sample batches with an odd number of samples, segmentation copy number was normalized to the median copy number of all samples in that segmentation. In sample batches with even numbers of samples, we normalized the copy number data to the [(N/2)+1]-th value in the ascending sort ordered list of copy numbers for that segmentation. In the case of a cohort containing two discrete copy number populations with equal numbers of samples, this ‘modified median’ normalization would normalize to the larger copy number population instead of normalizing to the mean of the two middle numbers, which would once again float between the discrete copy number populations and result in mean normalization skewing.

We implemented three filtering strategies to identify what we believe are high confidence CNV calls (**Fig. S9**). In regions of low raw coverage, relative fold coverage values can alter dramatically with small differences in sequencing coverage. This led to segmentations in regions with low raw sequencing coverage having much wider distributions in normalized coverage/cnv-copies, making it hard to distinguish discrete copy number populations from one another. For each segment, we calculated the average raw coverage across the segmentation region for all samples. If the segmentation average raw coverage was less than 20, we determined that segment to be un-callable, and it was filtered.

Similarly, segmentations with higher raw sequencing coverage but large standard deviations in segmentation copy number values for the sample cohort were also hard to confidently call as CNVs. To confidently distinguish which copy number population a sample belongs to, there needs to be a certain threshold of copy number standard deviation for samples in each population. To confidently call the copy number population identity for samples differing by one copy for 99.7 percent of samples, the standard deviation for each population has to be 1 divided by 6 standard deviations or 0.167. To estimate the standard deviation of the 2-copy population within each segmentation region, we calculated the standard deviation for all samples within the segmentation that had a copy number greater than 1.5 and less than or equal to 2.5. If the standard deviation for this ‘estimated 2-copy population’ was greater than or equal to 0.167, the segmentation was determined to be un-callable and was filtered. The implementation of our coverage and standard deviation filtering thresholds are shown in [Supplemental Figure S9].

Finally, we implemented a two-tiered threshold strategy for calling segments as being a CNV. First, a stringent threshold was implemented. A segment could not be called as being a CNV unless at least one sample had < 1.25 copies or > 2.75 copies. Once this threshold was met, the segment was identified as a CNV, and a less stringent threshold was applied to determine whether each sample had a CNV at that segment. This threshold categorized samples by a half-copy threshold, i.e. [>0.5 and <=1.5 copies] = 1-copy, [>1.5 and <=2.5 copies] = 2-copies, [>2.5 and <=3.5 copies] = 3-copies, etc. This two-tiered system protects against calling a rare CNV because of one sample having a borderline copy number value, but it also adapts to polymorphic regions by being more forgiving to calling samples as having the CNV once the segment has been identified as a CNV by meeting the stricter threshold.

For two of our capture batches, samples were only called as being 0, 2, or 4 copy populations for a polymorphic CNV in the *PSTPIP1* capture region (**Supplementary Table ST6**). We interpreted these populations as actually being 0, 1, and 2 copies, respectively, which would maintain the relative coverage relationship. This would be consistent with a matching deletion in the gnomAD structural variation database. When this assumption is made, over half of the samples in these capture batches had either a heterozygous deletion or a homozygous deletion. This large proportion of deletions in the cohort would dramatically skew the average coverage in the region to a level that would make an actual 2-copy population appear to be twice the coverage of the mean, consistent with calling the 2-copy population as a 4-copy population. For this reason, we believe that we have made a reasonable assumption in converting the 0, 2, 4 copy populations to 0, 1, and 2 copy populations. This highlights a weakness of relative coverage methods in general, as an overabundance of common polymorphic CNVs would skew the reference ratio and therefore skew the resulting copy number estimates.

After CNV analyses were performed on each capture batch separately, CNV summary statistics for all capture batches were combined. If a CNV was observed in one cohort but not another, all samples in the cohort in which the CNV was not observed were recorded as having two copies. If the genomic windows for a CNV called in two capture batches overlapped in 90% of the window, the CNV calls were assumed to be the same CNV; summary statistics were combined and one genomic window was chosen to represent all of the data. To evaluate the effectiveness of our targeted capture CNV analysis, we compared our targeted capture CNV results to CNV identified in the gnomAD structural variant database. This was done using the gnomAD Structural Variant database v2.1.

### 4.10. Generating PSTPIP1 tagged expression constructs

PSTPIP1-FLAG expression vectors were generated by cloning *PSTPIP1* into a pCMV2-FLAG expression vector. *PSTPIP1* was amplified from pDONR211-PSTPIP1 (HsCD00040737, Harvard PlasmID Repository). *PSTPIP1* point mutation constructs were generated using the Q5 Site-Directed Mutagenesis Kit (New England Biolabs E0554S). PSTPIP1-ΔSH3-FLAG expression vector was generated by amplifying the region of PSTPIP1 encoding amino acids 1-364 into pcmv2-FLAG. Primer sequences for cloning and mutagenesis are provided in **Supplementary Table ST7**.

### 4.10 Co-immunoprecipitation assays

We transfected 2 × 10^6^ HEK293 cells with *PSTPIP1* expression plasmids using TransIT-LT1 (Mirus MIR 2304) according to the manufacturer’s protocols. Twenty hours post-transfection, we transferred cells to 1.5 mL Eppendorf tubes and washed two times with 1-1.5 mL PBS without calcium and magnesium. We lysed cells by rotating in 400 µL Co-IP lysis buffer (50 mM Tris-HCl [pH 7.5], 120 mM NaCl, 3.0 mM EDTA, 1.0% TritonX-100, 1.5% Protease Inhibitor Cocktail [MilliporeSigma P8340]) at 4°C for 1 hour. We then pelleted cell debris centrifugation at 12,000 rcf for 15 minutes at 4°C and transferred the supernatants to new tubes. We washed 20-25 µL of anti-FLAG resin (MilliporeSigma F2426) per co-immunoprecipitation condition with 3X volumes of Co-IP buffer (50 mM Tris-HCl [pH 7.5], 120 mM NaCl, 3.0 mM EDTA, 1.0% TritonX-100). Each wash was rotated for at least 5 minutes at 4°C, and then we centrifuged the resin at 3000 rcf for 30 seconds at 4°C. We aliquotted the affinity resin into separate tubes for each co-immunoprecipitation condition. The resin was pelleted one more time and discarded the supernatant. We added 350 µL of HEK293 lysate to each tube. We rotated the resin/lysate mixes at 4°C for 6-8 hours. We washed the resins twice with 750 µL of Co-IP lysis buffer. For each wash, we rotated the resins for 10 minutes at 4°C, centrifuged at 3000 rcf for 30 seconds at 4°C to pellet resin, and discarded the supernatant. We then added THP-1 or Jurkat cell lysate was to resins as indicated below, and rotated at 4°C overnight.

#### THP-1 cell co-immunoprecipitation

We stimulated 6.75 × 10^6^ THP-1 cells per co-immunoprecipitation condition with 400 ng/mL phorbol 12-myristate 13-acetate (**PMA**) for 30 minutes on non-tissue culture treated plates. We collected the THP-1 cells and centrifuged at them 300 rcf for 5 minutes at 4°C. We washed cells two times with ice-cold PBS without calcium and magnesium. We distributed cells into 1.5 mL Eppendorf tubes and pelleted one more time. We aspirated the PBS supernatant and resuspended the cells in a combined total of 6 mL Co-IP lysis buffer per tube. We lysed cells by rotating for ∼4 hours at 4°C. We pelleted the cell debris by centrifugation at 12,000 rcf for 15 minutes at 4°C and pooled the lysate supernatants. We added 560 µL (∼6.3 × 10^6^ cells) of lysate to each co-immunoprecipitation condition.

We washed overnight co-immunoprecipitation affinity resins two times in 750 µL Co-IP lysis buffer, after which the Co-IP lysis buffer was discarded and affinity resins were resuspended in 50 µL of 3XFLAG peptide (300 ng/µL; MilliporeSigma F4799). We incubated affinity resins on ice for ∼2 hours with intermittent agitation by hand to elute off bound protein, after which we pelleted the resins one final time. We transferred the eluted supernatants to new tubes.

### 4.11 Antibodies

Anti-FLAG (MilliporeSigma A8592), anti-PTP-PEST (Cell Signaling D4W7W), and anti-WASp (Invitrogen PA5-37303). Raw western images can be found at https://figshare.com/projects/2022_hidradenitis_suppurativa_PSTPIP1_missense_enrichment/139684.

## Data Availability

The raw sequencing data was derived from samples without broad consent for genetic data sharing. The associated analysis code is publicly available through the Roberson Lab GitHub repository.

https://github.com/RobersonLab/2022_hs_pstpip1

https://figshare.com/projects/2022_hidradenitis_suppurativa_PSTPIP1_missense_enrichment/139684

## 5. Acknowledgements

We would like to thank the Evan Eichler lab for their generous gift of many of the fosmids used for this capture. DJM-H was supported by NIH training grant T32-AR007279-36. This work was supported in part by the Washington University in St. Louis Institute of Clinical and Translational Sciences (EDOR; #UL1-TR000448) and The Ohio State University Center for Clinical and Translational Science (BK, JK; #UL1-TR001070). Parts of this work were supported by the WUSTL Rheumatic Diseases Research Resource-based Center (P30-AR073752). Sequencing for this project was performed at the GTAC@MGI sequencing core, partially supported by NCI grant #P30 CA91842 and ICTS/CTSA grant #UL1 TR000448. Some of the analyses were performed on the Center for High-Performance Computer cluster at Washington University, which was partially provided through NIH grant #S10-OD018091. The content is solely the responsibility of the authors and does not necessarily represent the official views of the National Institutes of Health.

