## Supplemental information for "Rare missense variants in the SH3 domain of *PSTPIP1* are associated with hidradenitis suppurativa"

### 7. Supplementary Materials

#### 7.1. Supplemental table legends

**Table ST1 – Targeted capture regions and coverage.**

Targeted capture sequencing was performed using the listed fosmids and BACs to generate capture probes. Genomic start and stop sites for each fosmid are denoted for the human genome build GRCh37/hg19. Average fold coverage across the fosmid regions was calculated for each individual in our HS patient cohort. Mean Coverage in the table represents the average fosmid wide coverage of the entire patient cohort.

**Table ST2 – Transcripts use for burden assessment.**

Information for the transcripts used for assessing burden. Gene column contains the gene or gene region tested. The amino acid start and end, along with length correspond to either the whole protein or the region analyzed.

**Tables ST3-5 – List of all coding sequence variants detected**

Variants observed in targeted capture of HS cohort that alters amino acids (**ST3**), cause synonymous changes (**ST4**), or affects non-coding sequence (**ST5**). All reported variants fell within the first and last base of exonic regions of targeted genes. Genomic positions are denoted for human genome build GRCh37/hg19. REF = reference allele. ALT = observed variant. REF and ALT indicate a genomic base change with respect to the positive strand. AAF = alternate allele frequency in gnomAD sub-populations. A value of NA indicates that the variant wasn't observed in the sub-population. In **ST5**, the SAS sub-population didn't include genomic sequencing. An NA, in this case, could mean that the variant wasn't observed or (for deep intronic variants) the status is unknown. gnomAD subpopulation abbreviations: AFR – African / African American, AMR - Admixed American, EAS - East Asian, NFE - Non-Finnish European, SAS - South Asian.

**Table ST6 – *PSTPIP1* polymorphic CNV copy number correction summary**

This table shows the distribution of samples with the indicated copy numbers for the PSTPIP1 CNV. For Batch 1 and Batch 3 samples, distributions are shown before and after copy number correction.

**Table ST7 – Primers and oligos used in the study**

Primers are shown along with their name and the sequence. An 'X' in primer sequences indicates an equal mix of ACGT nucleotides for generating combinatorial indexes. A 'P' indicates a phosphothioate bond to increase the affinity of overhang bonds in Illumina Y adapters.

**Table ST8 – Number of identified variants in our cohort and gnomAD for the capture region**

Shown is a summary of the variants detected in the current HS cohort and the gnomAD data release. Synonymous variants resulted in the same amino acid. Protein-affecting variants altered amino acids (including premature stops). Non-coding variants did not overlap with the coding sequence. The majority of detected variants were intronic or intergenic.

**Table ST9 – Synonymous variant increased burden test**

Though more difficult to interpret, synonymous variants can have a functional impact through alteration of sequence motifs, reduction of protein half-life through non-optimal codon usage, or alteration of splice usage. We therefore also tested for increased burden of synonymous variants for each of our candidate genes and protein domains of interest. In the gene column, ECD = extracellular domain and NICD = Notch intracellular domain. The observed alt alleles are the count of the number of synonymous **alleles** observed in our data. The mean and standard deviation (SD) are derived from the control population simulations. Raw P-values are directly from the Poisson test, and the adjusted P-values are after Bonferroni correction. No gene had a significantly increased burden. A one-sided test was used but had a two-sided test been appropriate then it is likely that *MAML3* would have had fewer synonymous variants than expected. This was also the gene with many polyQ tract indels detected. We hypothesize that the difference between our results and gnomAD lies somewhere in the data processing decisions or library creation protocols.

#### 7.2. Supplemental figures

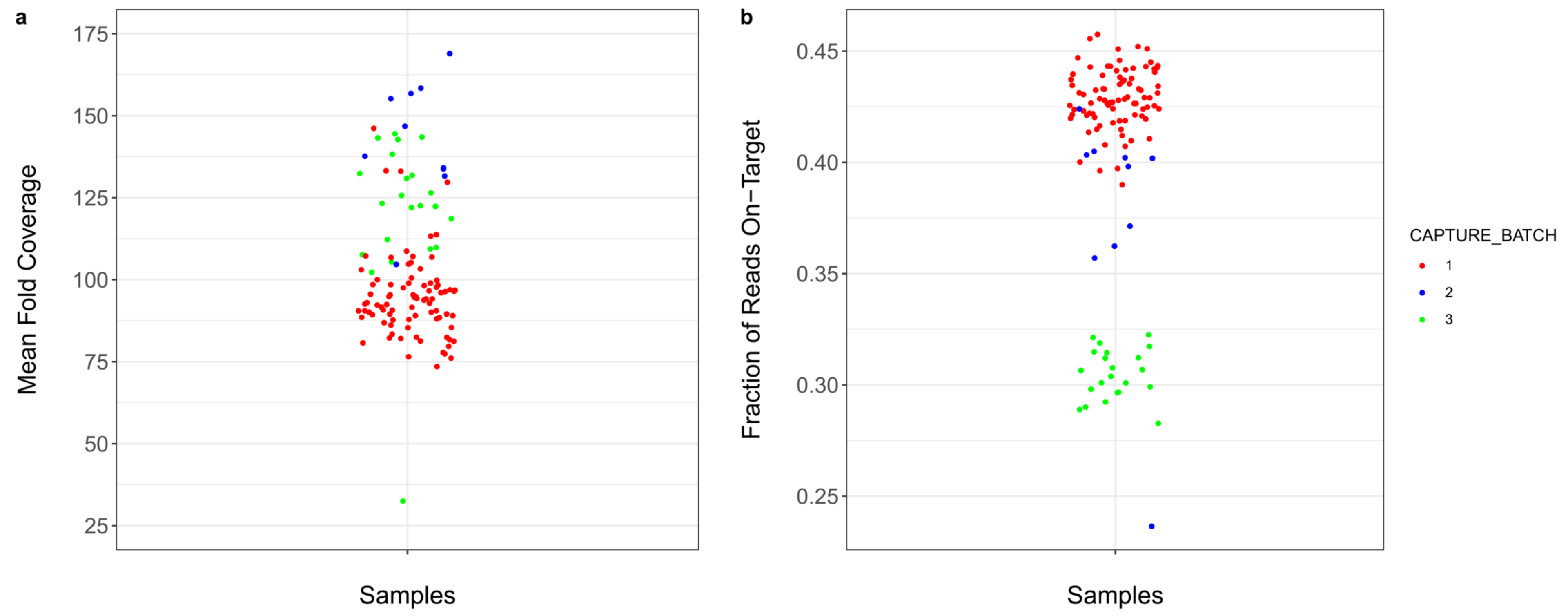


**Fig. S1 – Targeted capture performance by batch**
**a.** Average sequencing fold coverage over targeted capture regions for each individual in cohort. Almost all samples had at least 75X average coverage. **b.** The fraction of sequenced reads for each individual that overlapped with a targeted capture region by at least one nucleotide. The targeted capture region covered 2.57 megabases, making it about 0.08% of the GRCh37 genome. The worse performing sample had ~18% on-target and enrichment of approximately 225-fold. Most samples were even more efficient, supporting that the method works well.

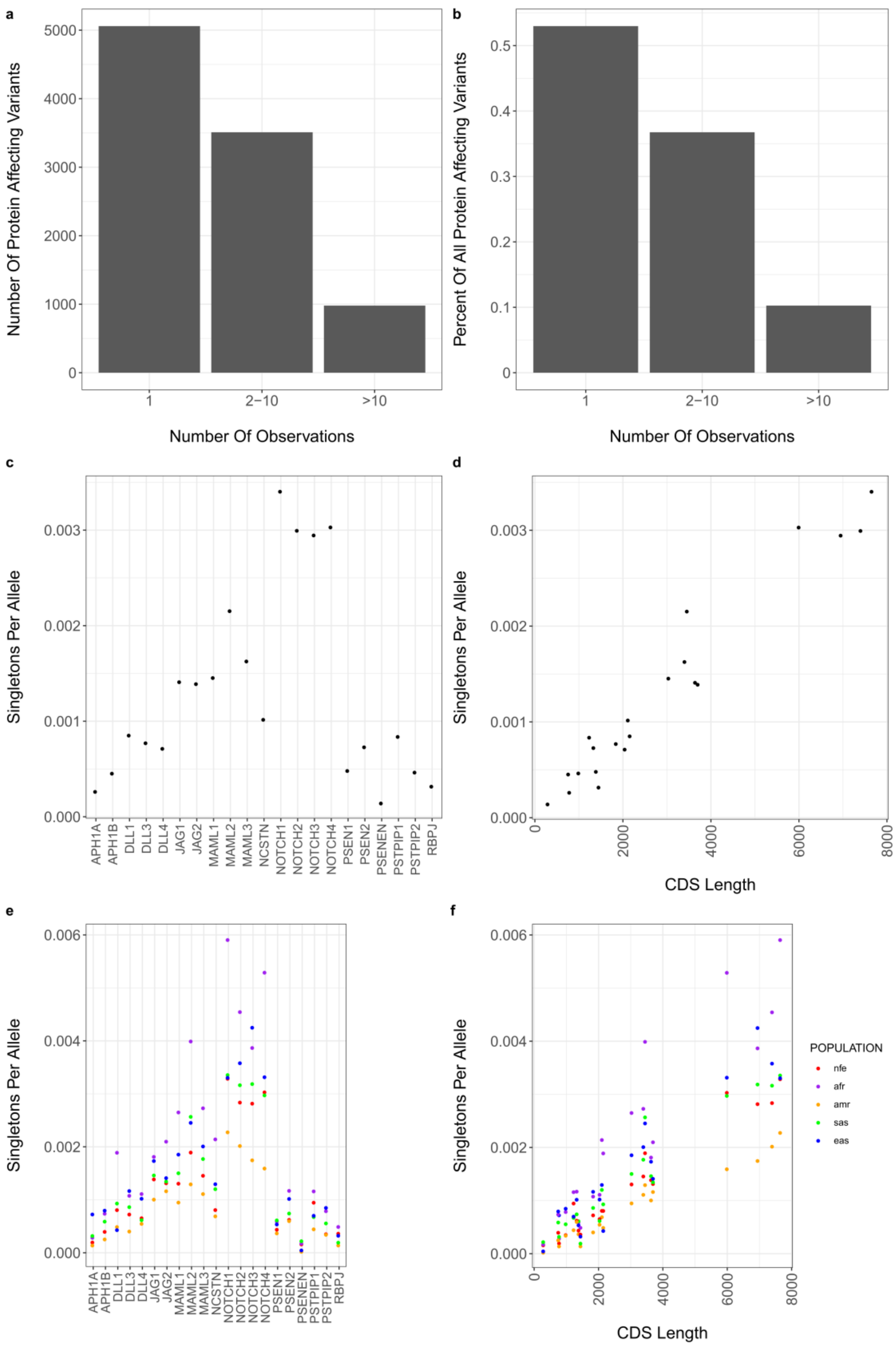


**Fig. S2 – Gene-specific singleton rates**

**a**, **b.** The majority of the gnomAD protein affecting variants in the targeted genes are variants for which only one allele has been observed. **c**. average gene-specific missense singleton frequencies. **d**. average gene-specific singleton frequencies are directly correlated with gene coding sequence length. **e**, **f**. gene-specific singleton frequencies were similar between geographic subpopulations.

*African*/African *American* – purple, *Admixed American* – yellow, *East Asian* – blue, *Non-Finnish European* – red, *South Asian* – green.

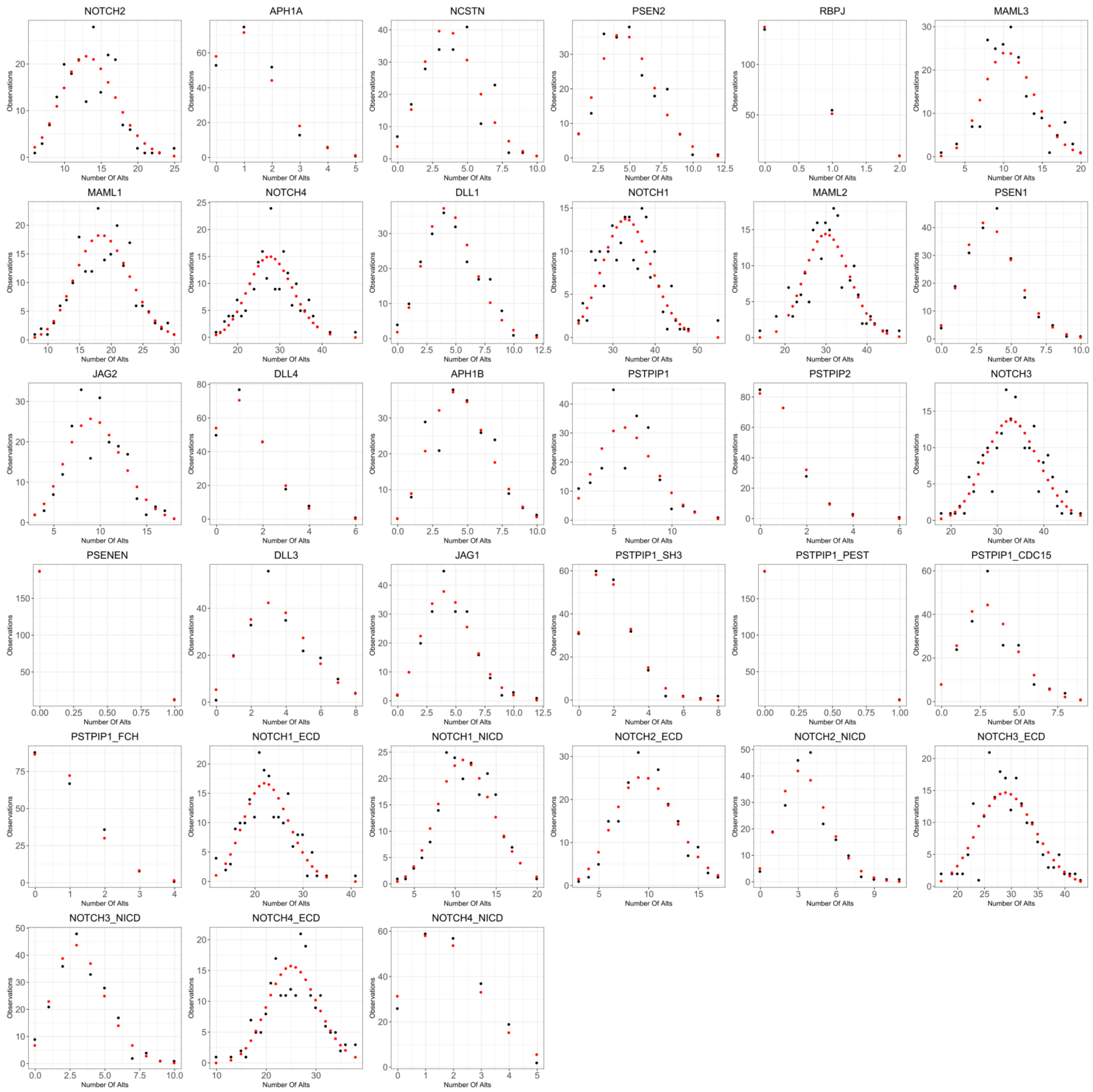

**Fig. S3 – Allele simulation outcomes compared to Poisson distribution**

We wanted to confirm the simulated outcomes followed a Poisson-like process. Each plot is a single gene or protein domain of a gene. The x-axis is the number of rare variants observed and the y-axis is a count of the number of times that outcome was observed. The red dots are the Poisson function using the mean number of alternative alleles as the lambda parameter, and the black dots are the individual simulation results. Overall, the simulations follow the expected distribution well.

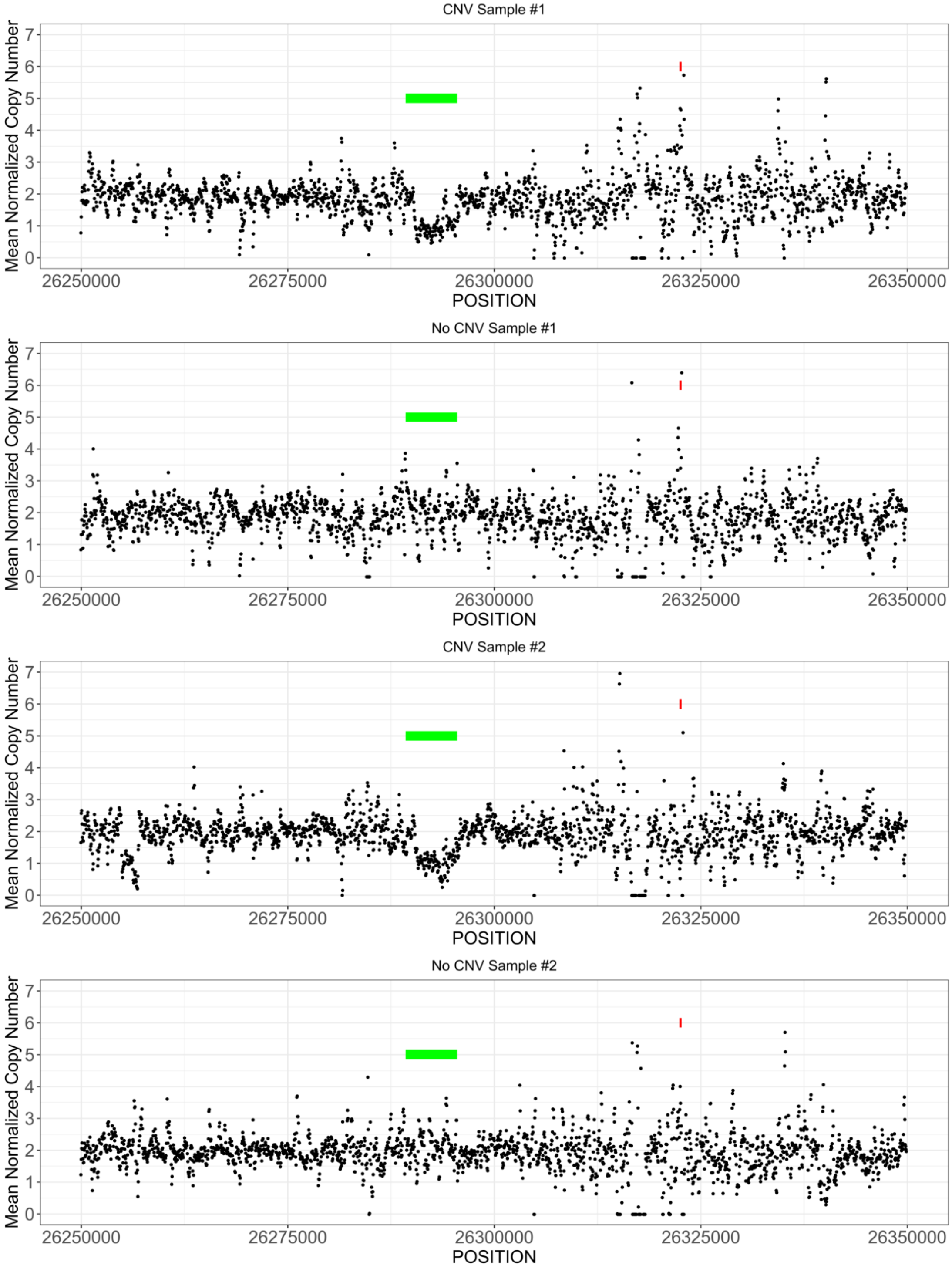


**Fig. S4 – *RBPJ* polymorphic CNV**

The x-axis shows the genomic location around the gene of interest. The y-axis is normalized copy number. Each dot represents the sliding window-smoothed estimated copy number of an individual point. Red bars indicate gene exons. Green bars indicated the CNV location.

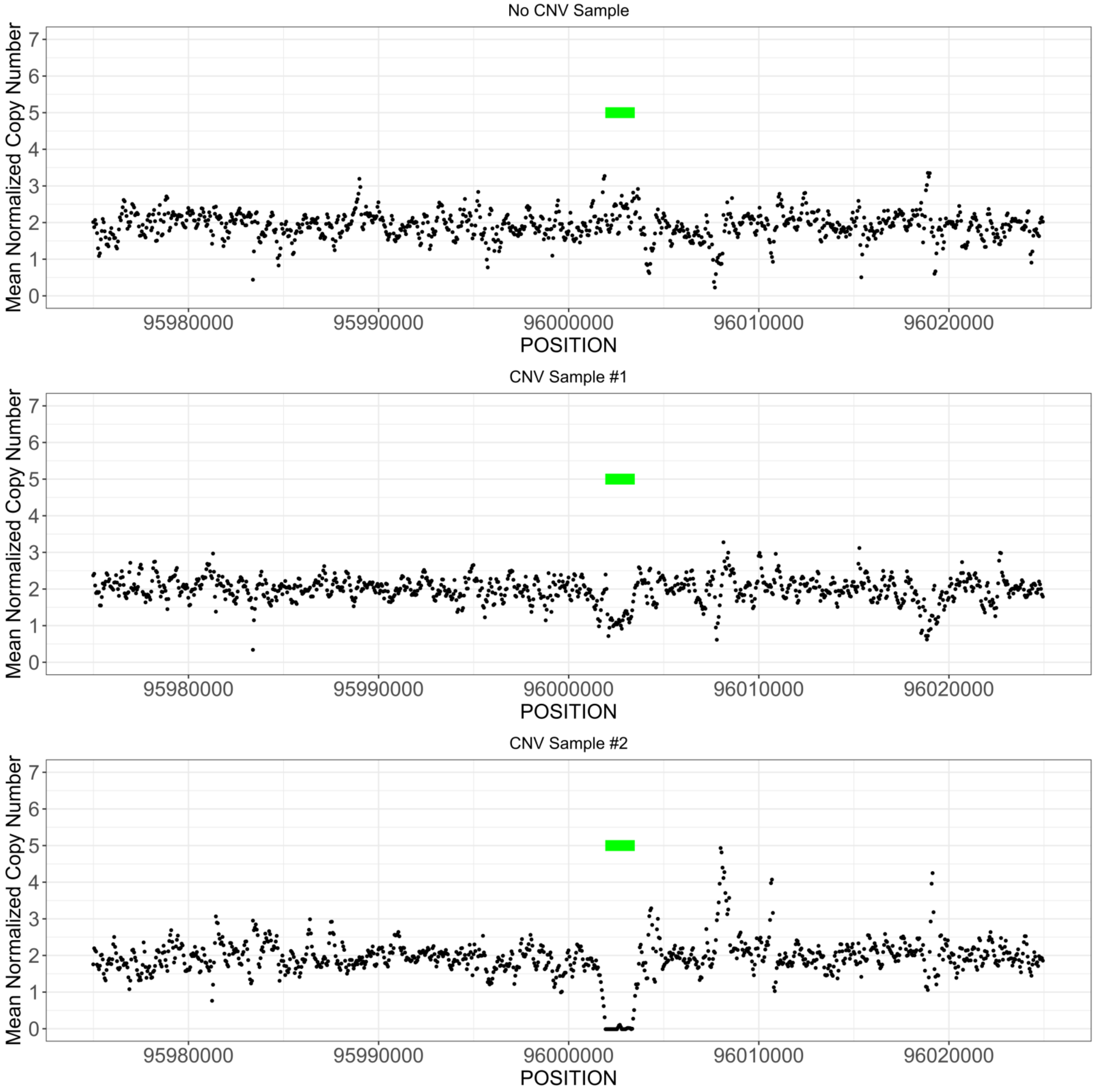


**Fig. S5 *MAML2* polymorphic CNV**
The x-axis shows the genomic location around the gene of interest. The y-axis is normalized copy number. Each dot represents the sliding window-smoothed estimated copy number of an individual point. Green bars indicated the CNV location. No exons are visible in this viewing window.

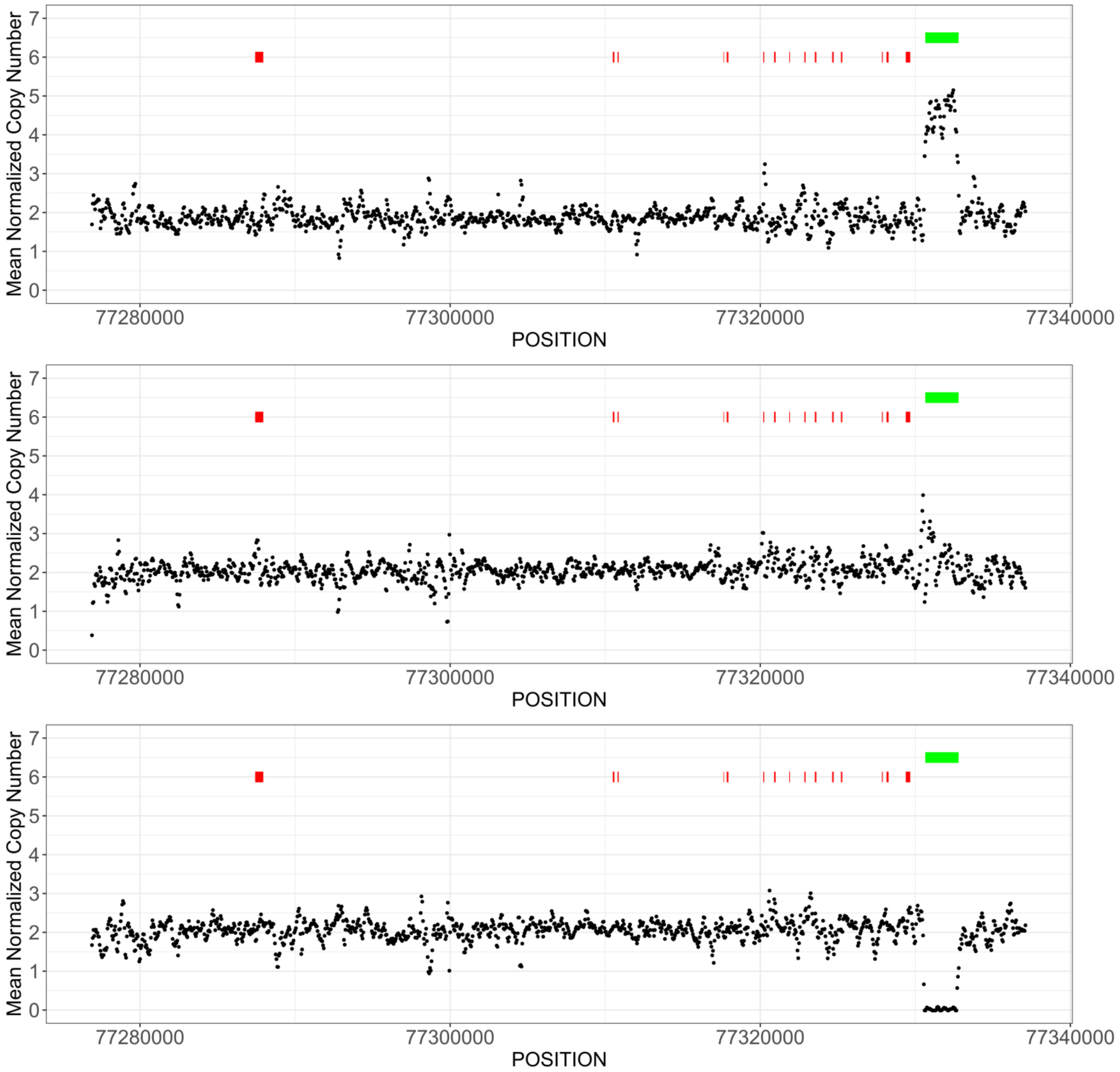


**Fig. S6 – *PSTPIP1* polymorphic CNV**
The x-axis shows the genomic location around the gene of interest. The y-axis is the normalized copy number. Each dot represents the sliding window-smoothed estimated copy number of an individual point. Red bars indicate gene exons. Green bars indicated the CNV location. Representatives were plotted for one of each of the distinct copy number populations observed. These copy number populations appear to be 0, 2, and 4 copies. However, we have interpreted the populations as 0, 1, and 2 copies. We believe that the high prevalence of the deletion (AAF > 0.5) caused mean normalized skewing to make the 1-copy population to be the average and thus interpreted as 2-copies in our mean normalization analysis (see Methods).

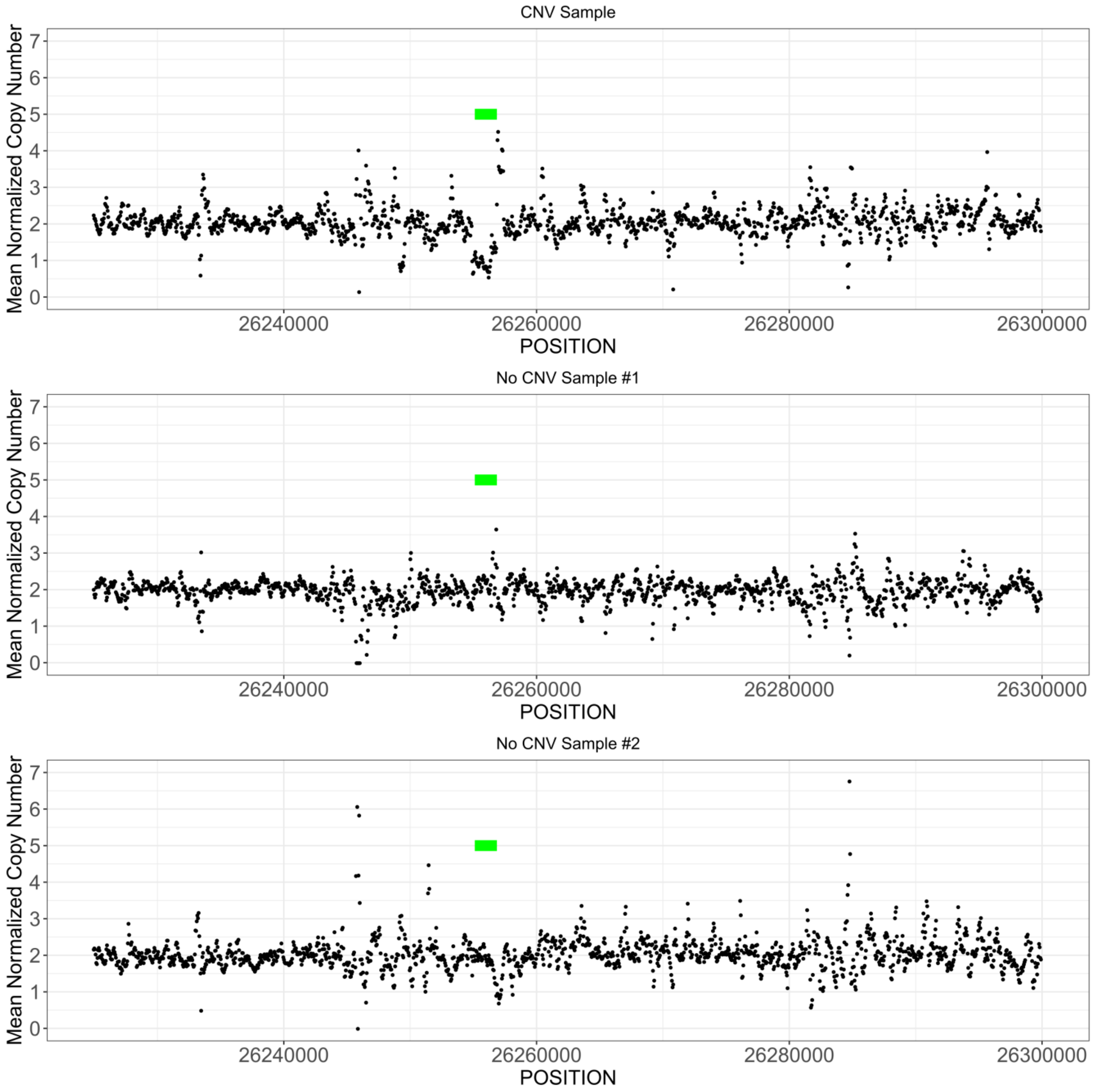


**Fig. S7 – *RBPJ* rare CNV**
The x-axis shows the genomic location around the gene of interest. The y-axis is the normalized copy number. Each dot represents the sliding window-smoothed estimated copy number of an individual point. Green bars indicated the CNV location. No exons are visible in this viewing window.

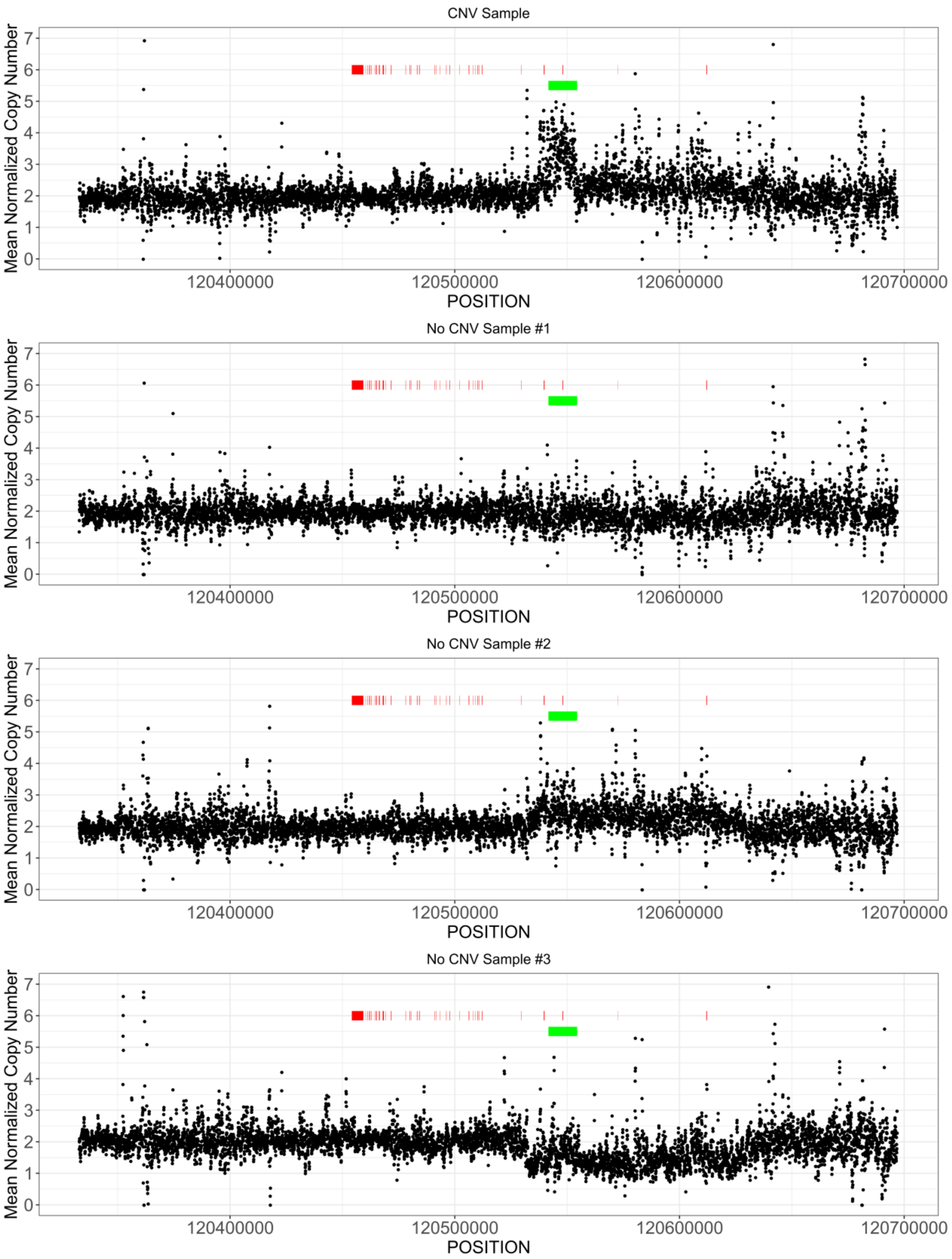


**Fig. S8 – *NOTCH2* rare CNV**

The x-axis shows the genomic location around the gene of interest. The y-axis is the normalized copy number. Each dot represents the sliding window-smoothed estimated copy number of an individual point. Red bars indicate gene exons. Green bars indicated the CNV location. In this case, the potential amplification overlaps an exon of the gene. If this is a true amplification it would be difficult to guess the exact impact on protein function and stability.

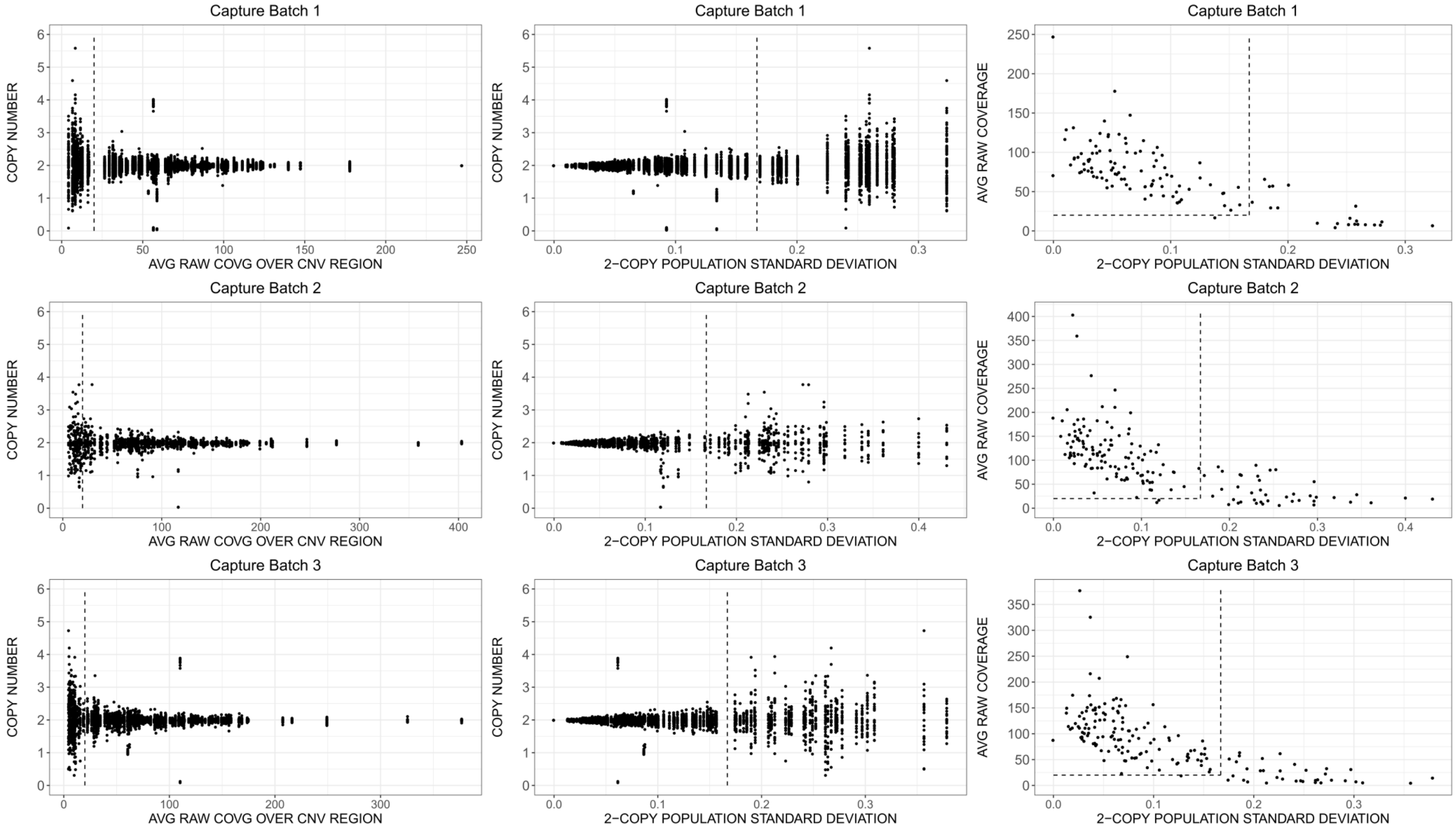


**Fig. S9 – CNV quality control filtering**

Plots showing the CNV filtering strategy described in the methods section. *Left column and middle column*, plots for each capture batch cohort showing the copy number for each sample in each segmentation as a function of the average raw coverage of the cohort across the segmentation or as a function of the standard deviation of all samples with copy number >1.5 and <=2.5 within the segmentation. *Right column*, plots for each capture batch cohort showing each segmentation as a function of the average raw coverage of the cohort across the segmentation or as a function of the standard deviation of all samples with copy number >1.5 and <=2.5 within the segmentation. Dashed lines indicate filter thresholds at average raw coverage = 20 and standard deviation = 0.167.

#### 7.3. Sequencing analysis details

For much of the genotyping, the small size of the capture panel made it worth limiting the genotyping and quality recalibration to only the fosmid targets. A BED formatted file listing the coordinates of the fosmid targets is referred to as Fosmid_Locations.bed. The number of threads used for multi-threaded operations is specified as "THREADS". Sample read group IDs (RGID) were derived from the flow-cell barcode + sample index. The de-identified sample name (RGSM). Training sites for GATK were from the GATK bundle (4.0.12.0). COHORT_SAMPLE_MAP is the tab-separated file describing the individual GVCF files and their RGSM, i.e. the first column is the RGSM and the second column is the path to the corresponding GVCF file.

##### 7.3.1. Cleaning with cutadapt

```
cutadapt \
-a AGATCGGAAGAGC \
-A AGATCGGAAGAGC \
-q 10 -m 30 -O 5 --match-read-wildcards
```

##### 7.3.2. Aligning with bwa

```
bwa aln \
-t THREADS \
/human_g1k_v37_decoy.fasta \
RGID_clean_1.fastq.gz 1>RGID_1.sai 2>RGID_1_aln.log

bwa aln \
-t THREADS \
human_g1k_v37_decoy.fasta \
RGID_clean_2.fastq.gz \
1>RGID_2.sai 2>RGID_2_aln.log

bwa sampe \
human_g1k_v37_decoy.fastq \
RGID_1.sai RGID_2.sai \
RGID_clean_1.fastq.gz RGID_clean_2.fastq.gz \
1>RGID.sam 2>RGID_sampe.log
```

##### 7.3.3. Pre-processing & GATK

```
gatk CleanSam \
--java-options “-Xmx11g” \
-MAX_RECORDS_IN_RAM 2750000 \
-INPUT RGID.sam \
-OUTPUT CLEAN.sam\

gatk AddOrReplaceReadGroups \
--java-options “-Xmx11g” \
-MAX_RECORDS_IN_RAM 2750000\
-SORT_ORDER coordinate \
-INPUT CLEAN.sam \
-OUTPUT RGSM_tagged_sorted.bam \
-RGID RGID \
-RGLB RGSM_1 \
-RGPL illumina \
-RGPU RGID \
-RGSM RGSM \

gatk MarkDuplicates \
--java-option “-Xmx14g” \
-MAX_RECORDS_IN_RAM 3500000 \
```

```
-INTPUT RGSM_tagged_sorted.bam \
-OUTPUT RGSM_tagged_sorted_dups_marked.bam \
-METRICS_FILE metrics.txt

gatk BuildBamIndex \
--java-options “-Xmx14g” \
-MAX_RECORDS_IN_RAM 3500000 \
-INTPUT RGSM_tagged_sorted_dups_marked.bam \
-OUTPUT RGSM_tagged_sorted_dups_marked.bai \

gatk BaseRecalibrator \
--java-options “-Xmx14g” \
-R human_g1k_v37_decoy.fasta \
-L Fosmid_Locations.bed \
-I RGSM_tagged_sorted_dups_marked.bam \
--known-sites dbsnp_138.b37.excluding_sites_after_129_sites.vcf \
--known-sites Mills_and_1000G_gold_standard.indels.b37_sites.vcf \
-O RGSM_bqsr.table

gatk BaseRecalibrator \
--java-options “-Xmx14g” \
-R human_g1k_v37_decoy.fasta \
-I RGSM_tagged_sorted_dups_marked.bam \
--bqsr-recal-file RGSM_bqsr.table \
-O RGID_BQSR.bam

BuildBamIndex \
--java-options “-Xmx14g” \
-MAX_RECORDS_IN_RAM 3500000 \
-INTPUT RGID_BQSR.bam \
-OUTPUT RGID_BQSR.bai
```

#### 7.3.4. Haplotype calling

```
gatk HaplotypeCaller \
--java-options “-Xmx14g” \
-R human_g1k_v37_decoy.fasta \
-L Fosmid_Locations.bed \
-I RGID_BQSR.bam \
--dbsnp dbsnp_138.b37_sites.vcf \
-O RGSM.gvcf \
--emitRefConfidence GVCF

gatk GenomicsDBImport \
--java-options “-Xmx19g” \
-L Fosmid_Locations.bed \
--genomicsdb-workspace-path /path/to/my_db \
--batch-size 50 \
--sample-name-map COHORT_SAMPLE_MAP

gatk GenotypeGVCFs \
--java-options “-Xmx35g” \
-R human_g1k_v37_decoy.fasta \
-V gendb://my_db \
-O raw_genotypes.vcf
```

#### 7.3.5. Variant filtering and merging

##### 7.3.5.1. SNPs only

```
gatk SelectVariants \
--java-options “-Xmx19g” \
-R human_g1k_v37_decoy.fasta \
-V raw_genotypes.vcf \
--select-type-to-include SNP \
-O snp_indels.vcf

gatk VariantFiltration \
--java-options “-Xmx19g” \
-R human_g1k_v37_decoy.fasta \
-V raw_snps.vcf \
-O marked_snps.vcf \
--filter-name "QD" \
--filter-expression 'QD < 2.0' \
--filter-name “MQ" \
--filter-expression 'MQ < 40.0' \
--filter-name "FS" \
--filter-expression 'FS > 60.0' \
--filter-name "SOR" \
--filter-expression 'SOR > 3.0' \
--filter-name "MQRankSum" \
--filter-expression 'MQRankSum < -12.5' \
--filter-name "ReadPosRankSum" \
--filter-expression 'ReadPosRankSum < -8.0'

gatk SelectVariants \
--java-options “-Xmx19g” \
-R human_g1k_v37_decoy.fasta \
-V marked_snps.vcf \
--exclude-filtered \
-O filtered_snps.vcf
```

##### 7.3.5.2. Indels only

```
gatk SelectVariants \
--java-options “-Xmx19g” \
-R human_g1k_v37_decoy.fasta \
-V raw_genotypes.vcf \
--select-type-to-include INDEL \
-O raw_indels.vcf

gatk VariantFiltration \
--java-options “-Xmx19g” \
-R human_g1k_v37_decoy.fasta \
-V raw_indels.vcf \
-O marked_indels.vcf \
--filter-name "QD" \
--filter-expression 'QD < 2.0' \
--filter-name "ReadPosRankSum" \
--filter-expression 'ReadPosRankSum < -20.0' \
--filter-name "Inbreeding" \
--filter-expression 'InbreedingCoeff < -0.8' \
--filter-name "FS" \
--filter-expression 'FS > 200.0' \
--filter-name "SOR" \
```

```
--filter-expression 'SOR > 10.0'

gatk SelectVariants \
--java-options “-Xmx19g” \
-R human_g1k_v37_decoy.fasta \
-V marked_indels.vcf \
--exclude-filtered \
-O filtered_indels.vcf
```

##### 7.3.5.3. Merging filtered data

```
gatk SortVcf \
-I filtered_snps.vcf \
-I filtered_indels.vcf \
-O filtered_variants.vcf
```
